# Blood-based multivariate methylation risk score for cognitive impairment and dementia

**DOI:** 10.1101/2023.09.27.23296143

**Authors:** Jarno Koetsier, Rachel Cavill, Rick Reijnders, Joshua Harvey, Kay Deckers, Sebastian Köhler, Lars Eijssen, Rebecca G. Smith, Adam R. Smith, Joe Burrage, Emma M. Walker, Gemma Shireby, Eilis Hannon, Emma Dempster, Tim Frayling, Jonathan Mill, Valerija Dobricic, Yasmine Sommerer, Peter Johannsen, Michael Wittig, Andre Franke, Rik Vandenberghe, Jolien Schaeverbeke, Yvonne Freund-Levi, Lutz Frölich, Philip Scheltens, Charlotte Teunissen, Giovanni Frisoni, Olivier Blin, Jill Richardson, Régis Bordet, Sebastiaan Engelborghs, Ellen de Roeck, Pablo Martinez-Lage, Mikel Tainta, Alberto Lleó, Isabel Sala, Julius Popp, Gwedoline Peyratout, Frans Verhey, Magda Tsolaki, Ulf Andreasson, Kaj Blennow, Henrik Zetterberg, Johannes Streffer, Stephanie J. B. Vos, Simon Lovestone, Pieter-Jelle Visser, Lars Bertram, Katie Lunnon, Ehsan Pishva

**Author notes:** Corresponding author: Ehsan Pishva, Maastricht University, 6200 MD Maastricht, The Netherlands.

## Abstract

**INTRODUCTION:** Given the established association between DNA methylation and the pathophysiology of dementia and its plausible role as a molecular mediator of lifestyle and environment, blood-derived DNA methylation data could enable early detection of dementia risk.

**METHODS:** In conjunction with an extensive array of machine learning techniques, we employed whole blood genome-wide DNA methylation data as a surrogate for 14 modifiable and non-modifiable factors in the assessment of dementia risk in two independent cohorts of Alzheimer’s disease (AD) and Parkinson’s disease (PD).

**RESULTS:** We established a multivariate methylation risk score (MMRS) to identify the status of mild cognitive impairment (MCI) cross-sectionally, independent of age and sex. We further demonstrated significant predictive capability of this score for the prospective onset of cognitive decline in AD and PD.

**DISCUSSION:** Our work shows the potential of employing blood-derived DNA methylation data in the assessment of dementia risk.

## 1. BACKGROUND

As a result of population aging, the global number of dementia cases has drastically increased over the past decade and is estimated to keep increasing to more than 150 million patients worldwide by 2050 [1]. Despite the seemingly ever-increasing global burden, there is currently no availability of drugs or other treatment options that can halt or fully reverse the cognitive decline in dementia [2]. This emphasizes the importance of pre-clinical prevention strategies that require the identification of persons at risk of developing dementia before the onset of irreversible cognitive decline.

Over the past decades, the most precise models for predicting dementia and Alzheimer’s disease (AD) have relied on molecular information derived from cerebrospinal fluid (CSF) and neuroimaging modalities [3, 4]. In addition to that, several studies have identified genetic and environmental (lifestyle-related) factors that are associated with dementia risk, such as smoking [5], alcohol intake [6], plasma cholesterol levels [7], physical activity [8], education [9], and diet [10]. Accordingly, several of these unmodifiable and modifiable (lifestyle-related) dementia risk factors have been combined into a single score, such as the ‘*Cardiovascular Risk Factors, Aging, and Dementia’* (CAIDE) [11] and *‘Lifestyle for Brain Health’* (LIBRA) [12] scores, respectively. However, in recent years, there has been a significant shift towards prioritizing blood-based biomarkers for AD [13]. This is due to several factors, including the invasive nature of CSF sampling, the lack of objective quantitative assessment of lifestyle-related factors, and the high cost and limited availability of specialized neuroimaging facilities.

It has been well-established that epigenetic mechanisms and in particular, DNA methylation, are involved in the molecular pathology of various neurodegenerative diseases, including dementia [14]. DNA methylation is a molecular mechanism that can mediate the impact of lifestyle and environmental factors on the genome, regulating the expression of the genes [15]. Notably, prior research has demonstrated associations between peripheral DNA methylation patterns and risk factors for dementia, such as smoking [16], obesity [17], and blood pressure [18]. Moreover, recent comprehensive DNA methylation Quantitative Trait Locus (mQTL) analyses have confirmed a substantial genetic influence on methylation patterns [19]. This collective evidence positions DNA methylation as an intriguing molecular biomarker with the potential to capture both genetic and environmental information at the individual level. However, previous endeavors to establish blood-derived DNA methylation-based predictions for AD have encountered challenges in external validation, likely attributed to the heterogeneity of the disease among different cohorts [20].

Therefore, in this study, rather than adopting a direct approach to predict the risk of dementia using blood DNA methylation, we sought to utilize the large-scale nature of a general population cohort to first develop objective and precise DNA methylation-based prediction models for the CAIDE and LIBRA scores and multiple modifiable and non-modifiable dementia risk factors. Next, along with the epigenetic score for CAIDE and LIBRA (*i.e.,* epi-CAIDE and epi-LIBRA), we employed the methylation profile scores (MPSs) of the individual dementia risk factors to generate a multivariate methylation risk score (MMRS) for cognitive impairment and dementia.

## 2. METHODS

The applied methodology consists of four main steps: (1) the *model generation* by two distinct approaches using the DNA methylation data of the Exeter 10,000 project (EXTEND) [21] and the European Medical Information Framework for Alzheimer’s Disease (EMIF-AD) [22] cohorts (**Fig. 1**), (2) *model validation* in the independent test set of the EMIF-AD, Parkinson’s Progression Markers Initiative (PPMI) [23] and the Alzheimer’s Disease Neuroimaging Initiative (ADNI) [24] cohorts, (3) *model interpretation* in terms of variable importance, gene ontology (GO) overrepresentation analysis, and the influence of genetic variation, and (4) *model extension* by adding genetic and CSF biomarkers to the model.

**FIGURE 1.**
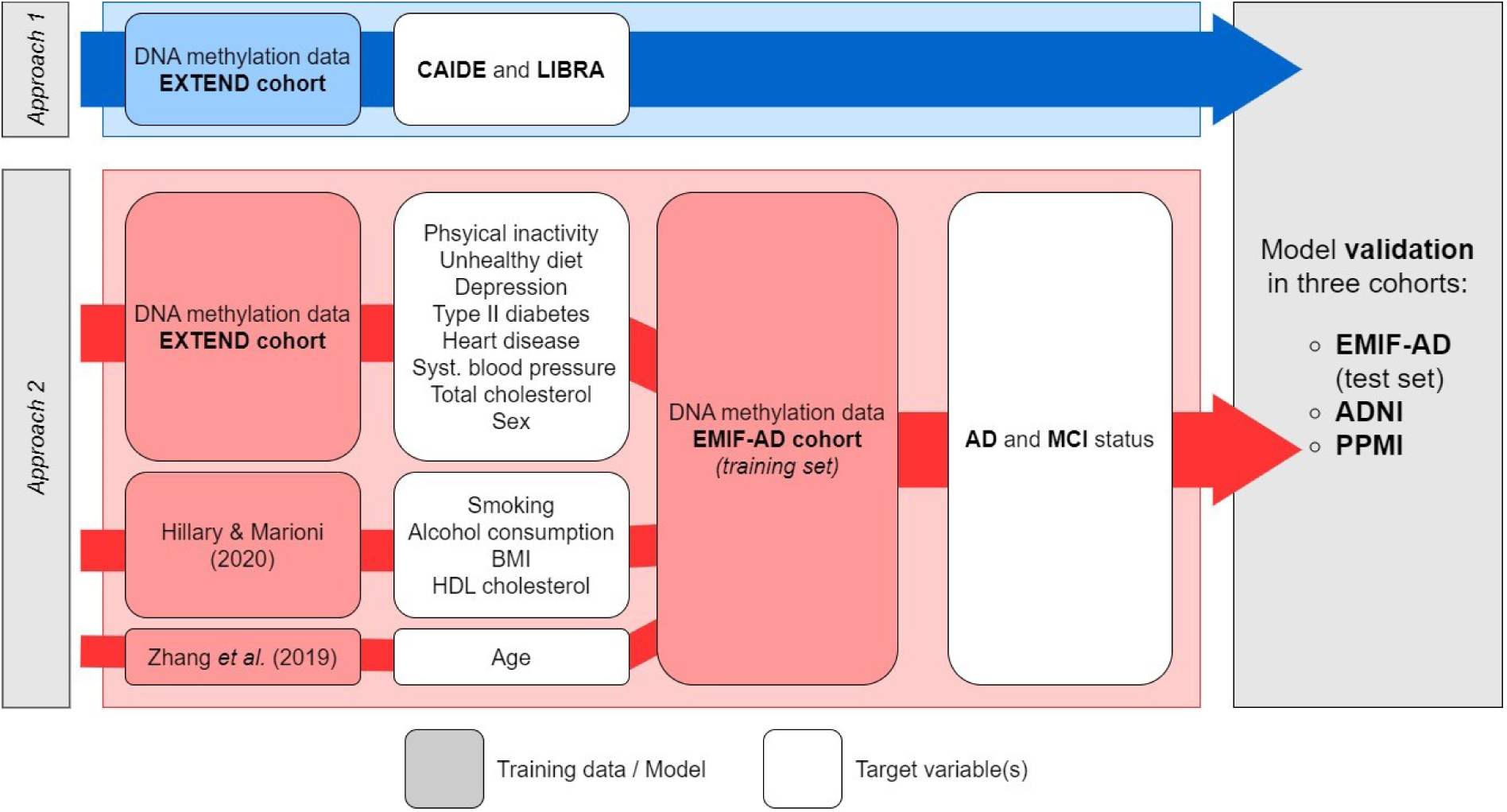
Overview of the model generation. The model generation workflow consists of model training in the EXTEND and EMIF-AD cohorts using two different approaches. In the first approach, models for the prediction of CAIDE and LIBRA were trained in the EXTEND cohort (*section 2.3.1*.). Furthermore, in approach 2, the EXTEND cohort was used to predict 14 known dementia risk factors. The predicted risk scores by these 14 models (*i.e.,* methylation profile scores; MPSs) were used as variables for the prediction of AD and MCI status in the training set of the EMIF-AD cohort (*section 2.3.2*.). Both approaches were evaluated in terms of AD and MCI classification performance in the independent test set in the EMIF-AD cohort. The model with the best performance was also validated in the PPMI and ADNI longitudinal cohorts (*section 2.4*.).

### 2.1. Study cohorts

#### 2.1.1. The EXTEND cohort

The EXTEND study is a National Institute for Health and Care Research (NIHR) funded project aiming to collect blood samples along with extensive health information from people with and without health issues [21]. In the current study, a subset of the EXTEND cohort, consisting of individuals with available genotyping and blood-derived DNA methylation data (n = 1076) was used. The dataset exclusively comprised phenotype data from individuals aged 40 to 75, denoted as the midlife age group.

#### 2.1.2. The EMIF-AD cohort

The EMIF-AD cohort is a research initiative focused on collecting and collating comprehensive medical and health-related information from individuals affected by AD and related cognitive disorders [22]. In this study, besides excluding the individuals outside the midlife age range (*i.e.,* age < 40 or age > 75), cognitively healthy individuals who were recorded to have converted to MCI or AD during the follow-up period (mean follow-up time ± sd ≈ 2.3 ± 1.2 years) were also excluded. This process resulted in a final dataset of 110 individuals with AD, 293 individuals with MCI, and 220 healthy controls with available blood-derived DNA methylation data, genotyping data, and CSF protein markers.

#### 2.1.3. The ADNI cohort

The ADNI cohort [24] contains genetic and blood-derived DNA methylation data of cognitively normal individuals as well as individuals diagnosed with MCI and dementia. In addition to the DNA methylation data, ADNI has also extensive individual-level information on various psychometric biomarkers measured at multiple time points. For the validation of the models, we used the baseline DNA methylation data of 223 midlife (40 ≤ age ≤ 75) individuals with a Mini Mental State Examination (MMSE) ≥ 26 (*i.e.,* cognitively intact) at baseline.

#### 2.1.4. The PPMI cohort

The Parkinson’s Progression Markers Initiative (PPMI) cohort [23] includes the blood-derived DNA methylation data of individuals recently diagnosed with Parkinson’s Disease (PD). Only samples from persons with 40 ≤ age ≤ 75 were included for the validation, resulting in 129 samples for which baseline DNA methylation data and the cognitive impairment outcome information were available. An overview (*i.e.,* description, sample size, and sex and age distribution) of the four cohorts used in the current research is provided in **Supplementary Table 1**.

### 2.2 Data preprocessing

#### 2.2.1. Clinical outcomes

The cognitive status of individuals in the EMIF-AD cohort was defined as described previously [22]. In summary, cognitively healthy individuals were defined by a normal neuropsychological assessment score. In nine of the EMIF-AD subcohorts, the MCI diagnosis was based on the criteria of Petersen [25], while for two subcohorts the Winblad *et al.* criteria [26] was used. Furthermore, AD diagnosis was defined based on the criteria of the National Institute of Neurological and Communicative Disorders and Stroke–Alzheimer’s Disease and Related Disorders Association (NINCDS-ADRDA) [27].

In the ADNI cohort, we only included the data of cognitively intact (*i.e.,* MMSE ≥ 26) individuals at baseline to perform the survival analysis based on the longitudinal outcome of different cognitive domains (*i.e.,* Alzheimer’s Disease Assessment Scale (ADAS), Rey’s Auditory Verbal Learning Test (RAVLT), Wechsler Logical Memory Delay (LDELTOTAL), Trail Making Test Part B Time (TRABSCOR), and MMSE scores) measured over 4 years. For the MMSE score we defined cognitive impairments by a MMSE < 24, while for the other cognitive outcomes, cognitive impairment status was defined by a score of either 2 SD below or above the mean of the control group, depending on the direction of the score (**Supplementary Table 2**).

Finally, in the PPMI cohort, dementia and MCI diagnosis was based on the Movement Disorders Society (MDS) recommended criteria [28, 29] as done previously [30]. MCI and dementia individuals who have been recorded to have reverted to a cognitively normal status (absence of MCI and dementia) were excluded from the analysis.

#### 2.2.2. Dementia risk factors

In the current study, the dementia risk factors were defined in the EXTEND cohort according to **Table 1**. Particularly, 15 dementia risk factors were used for the calculation of the CAIDE and LIBRA scores based on the previously established risk factor weights [12, 31] (**Supplementary Tables 3 and 4**). Additionally, because of the small number of kidney disease cases, only 14 dementia risk factors were used as target variables for the construction and/or validation of the DNA methylation-based risk factor models (*section 2.3.2*.).

**TABLE 1.** Risk factor definitions in the EXTEND cohort. The risk factors in the EXTEND cohort were used for the calculation of the CAIDE and LIBRA scores and as target variables for the training or validation of the DNA methylation-based risk factor models.

| **Risk factor** | **Definition** | **Type** | **Risk factor model \*** | **CAIDE †** | **LIBRA †** |
| --- | --- | --- | --- | --- | --- |
| *Low education* | A person is defined to be lowly educated if the individual has none of the following educational achievements:<br>1. College or university degree.<br>2. O level, GCSEs, or equivalent.<br>3. NVQ, HND, HNC, or equivalent.<br>4. A-level, AS-level, or equivalent.<br>5. CSEs or equivalent.<br>6. Other professional qualifications | Binary | Training in EXTEND | **X** |  |
| *Physical inactivity* | A person is defined to be physical inactive if self-reported to do exercise with increased pulse more than 2.5 hours per week. | Binary | Training in EXTEND | **X** | **X** |
| *Unhealthy diet* | Self-reported consumption of three or less fruits/vegetables per day | Binary | Training in EXTEND |  | **X** |
| *Depression* | Self-reported depression status | Binary | Training in EXTEND |  | **X** |
| *Type II diabetes* | Self-reported type II diabetes status | Binary | Training in EXTEND |  | **X** |
| *Heart disease* | Self-reported heart disease status | Binary | Training in EXTEND |  | **X** |
| *Sex* | Sex | Binary | Training in EXTEND | **X** |  |
| *Systolic blood pressure* | Mean systolic blood pressure (mmHg) | Continuous | Training in EXTEND |  | **X** |
| *Total cholesterol* | Log-transformed total serum cholesterol levels (mmol/L) | Continuous | Training in EXTEND | **X** |  |
| *Age* | Chronological age | Continuous | Zhang *et al.* (2019) model | **X** |  |
| *BMI* | Body mass index | Continuous | Hillary & Marioni (2020) model | **X** | **X** |
| *Smoking* | Self-reported current smoking status | Binary | Hillary & Marioni (2020) model |  | **X** |
| *HDL cholesterol* | Log-transformed serum HDL cholesterol levels (mmol/L) | Continuous | Hillary & Marioni (2020) model |  | **X** |
| *Alcohol intake* | High alcohol intake is defined by the following criteria:<br>1. Once a month, more than 10 alcoholic drinks per day.<br>2. 2-4 a month, 5 or more alcoholic drinks per day<br>3. 2-3 a week, 5 or more alcoholic drinks per day<br>4. 4 or more a week, 3 or more alcoholic drinks per day | Binary | Hillary & Marioni (2020) model |  | **X** |
| *Kidney disease ‡* | Self-reported chronic kidney disease status | Binary | NA ‡ |  | **X** |

\* This column indicates the DNA methylation-based model that was used for the prediction of the corresponding dementia risk factor (approach 2).
† This column indicates whether the corresponding risk factor was used in the calculation of the LIBRA and CAIDE score (approach 1). See Supplementary Tables 3 and 4 for the exact risk factor weights.
‡ No risk factor model was trained for kidney disease status due to the limited number of cases in the EXTEND cohort (*i.e.*, 17 cases and 1059 controls).

#### 2.2.3. Cerebral spinal fluid biomarkers

Three CSF biomarkers scores (amyloid-β (Aβ), phosphorylated tau (p-tau), and total tau (t-tau) z-scores) were defined in the EMIF-AD cohort as described previously [22]. In summary, the t-tau and p-tau z-scores were defined by their local p-tau levels as measured by center-specific ELISAs, standardized within assay according to mean and standard deviation of the healthy controls. Furthermore, the Aβ z-score were specified to be standardized scores for amyloid pathology. Specifically, this score was standardized within assay according to the mean and standard deviation of the control group and was based on CSF Aβ42/40 ratio, local CSF-Aβ42, and the standardized uptake value ratio (SUVR) on an amyloid PET scan.

#### 2.2.4. Whole blood DNA Methylation profiling

Sample filtering was performed as a first step of the DNA methylation data pre-processing pipeline and includes the removal of samples with a median bisulfite conversion rate below 80 percent, incorrect sex labels, and a low median (un)methylated intensity according the *minfi* package’s guidelines (*i.e.,* median log_2_ unmethylated intensity + median log_2_ methylated intensity ≤ 21) [32].

In the normalization procedure, the combination of Noob (*minfi* package (v1.46.0) [32]) and BMIQ (*wateRmelon* package (v2.6.0) [33]) normalization was applied. This pipeline has previously been shown to be a high-performing method for reducing type I/type II bias and enhancing reproducibility [34]. Furthermore, both Noob and BMIQ are within-sample normalization methods, which avoid information leakage from the training to the test set.

Before model training in the EXTEND cohort, previously reported cross-reactive probes [35, 36], probes with a detection P value > .01 in at least one sample, sex-chromosomal probes, and probes with single nucleotide polymorphisms (SNPs) at the single base extension and/or CpG interrogation site were removed. Next, density plots and principal component analysis (PCA) score plots were constructed to assess the quality of the pre-processing and to identify possible outliers. Lastly, β-values were converted to M-values to account for the inherently heteroscedastic nature of methylation data. The M-values were used for the subsequent feature selection and machine learning pipeline.

It should be noted that not all features that are incorporated in the generation of the risk factor models passed the described probe filtering steps in the other cohorts. Therefore, when applying the risk factor models in a different cohort, low-quality probes (*i.e.,* detection P value > .1) were imputed using the *imputePCA* function from the *missMDA* package (v1.8) [37]. This function uses a regularized iterative PCA algorithm to impute missing values. Specifically, this algorithm first imputes all missing values with the feature’s mean after which it iteratively performs PCA and imputes each missing value using the low-rank representation until convergence is reached (*i.e.,* a difference of less than 10^-6^ between iterations). The optimal number of principal components (PCs) for the low-rank representation was found by removing and imputing 100 random values and choosing the number of PCs that yields the lowest mean absolute error (MAE). This iterative PCA imputation method has previously been shown to be among the best-performing and computationally efficient algorithms for missing value imputation of DNA methylation data [38].

#### 2.2.5. Polygenic score generation

The genomics data from the EMIF-AD cohort was pre-processed as described previously [39]. In short, this pre-processing pipeline includes filtering of strand-ambiguous SNPs, aligning alleles to the human genome assembly GRCh37/hg19, phasing, imputation based on the HRC reference panel, pre- and post-imputation quality control (QC), and filtering of SNPs with a minor allele frequency (MAF) < 0.01. The genomics data of the EXTEND cohort underwent a similar pre-processing pipeline including SNP filtering and alignment using the HRC/100G imputation preparation and checking pipeline (v4.2.7) [40], imputation using the Michigan Imputation Server with the 1000Genomes reference panel (phase 3 v5 hg19, Population: EUR, Phasing: Eagle, R-squared filter: 0.3) [41], and post-imputation SNP filtering (MAF < 0.01, Hardy-Weinberg equilibrium (HWE) P value < 10^-4^, missing call rated > 0.1).

Subsequently, the LDAK tool (v5.2) [42] was applied to calculate the polygenic (risk) scores (PGSs) for 11 dementia risk factors as well as AD status (including the APOE region) using the HapMap reference panel and the summary statistics of 12 genome-wide association studies (GWASs) [43–53] (**Supplementary Table 5**). In short, LDAK splits the summary statistics in pseudo training and test summary statistics and then uses a variational Bayes approach to estimate the regression coefficients of the SNPs [42]. For the hyperparameter optimization, multiple models are trained and evaluated on the test summary statistics for different combinations of prior distribution parameters. This methodology is available for six model types (*i.e.,* bayesR, bayesR-shrink, lasso, lasso-sparse, ridge, and bolt regression models) that differ in the form of the prior distribution for the SNP effect sizes. As the bayesR is the recommended method for PGS generation by the developers of LDAK, all PGSs were generated using the bayesR approach.

### 2.3. Model generation

The model generation consists of two approaches; the prediction of the CAIDE and LIBRA scores in the EXTEND cohort as well as the prediction of MCI and AD status in the EMIF-AD cohort by 14 methylation profile scores of dementia risk factors (Fig. 1).

#### 2.3.1. Approach 1: Generation of methylation risk scores for CAIDE and LIBRA

In the first approach, we aimed at constructing an epigenetic model for the prediction of the LIBRA score (*i.e.,* epi-LIBRA model) and CAIDE score (*i.e.,* epi-CAIDE model). However, before constructing these models, we evaluated multiple supervised and unsupervised feature selection methods to investigate which method is suitable for reducing the large dimensionality of the DNA methylation data and found superior performance of the “correlation-based feature selection method” (see **Supplementary Text 1** for a more detailed description of the applied methodology and results). In short, in the correlation-based feature selection method, the 10,000 CpGs that have the highest absolute Spearman correlation coefficient with the target variable in the training set were selected for model training.

Hence, for the prediction of both the CAIDE and LIBRA scores, we trained an ElasticNet and Random Forest model on the 10,000 features selected by correlation-based feature selection as well as an ElasticNet model trained on all CpGs that passed QC (**Table 2**). Accordingly, 5-repeated 5-fold cross-validation was applied to find the optimal hyperparameter values that yield the minimal MAE (the searched hyperparameter space is shown in **Supplementary Table 6**).

**TABLE 2.** The applied feature selection and machine learning methods. For the prediction of nine dementia risk factors and the CAIDE and LIBRA scores in the EXTEND cohort, different combinations of feature selection and machine learning methods were used.

| Feature selection method | Machine learning method | CAIDE/LIBRA | Dementia risk factors |
| --- | --- | --- | --- |
| *None* | *ElasticNet* | **x** | **x** |
| *Correlation-based \** | *Random Forest* | **x** | **x** |
| *Correlation-based \** | *ElasticNet* | **x** | **x** |
| *Literature-based †* | *Random Forest* |  | **x** |
| *Literature-based †* | *ElasticNet* |  | **x** |

\* In the *correlation-based feature selection* method, the 10,000 CpGs that have the highest absolute Spearman correlation coefficient with the target variable (*i.e.*, predicted dementia risk factor) in the training set were selected for model training (*Supplementary Text 1*).
† In the *literature-based feature selection* method, the CpGs that reached genome-wide significance in a previously performed epigenome-wide association study (*Supplementary Table 7*) for the corresponding risk factor were selected for model training.

#### 2.3.2. Approach 2: Generation of multivariate methylation risk scores

For the second approach, we aimed at predicting 14 known dementia risk factors that are included in the CAIDE and/or LIBRA scoring systems (**Table 1**). Specifically, for the prediction of smoking, alcohol consumption, HDL cholesterol, and BMI, the corresponding DNA methylation-based models from Hillary and Marioni (2020) [54] were used, while the epigenetic clock model from Zhang *et al.* (2019) [55] was used for the prediction of age. For each of the nine remaining risk factors (*i.e.,* low education, physical inactivity, unhealthy diet, depression, type II diabetes, heart disease, sex, systolic blood pressure, and total cholesterol), five models were trained in the EXTEND cohort corresponding to different combinations of feature selection and machine learning algorithms (**Table 2**). These include an ElasticNet model without prior feature selection, an ElasticNet and Random Forest model with correlation-based feature selection, as well as an ElasticNet and Random Forest model trained on the CpGs that reached genome-wide significance in previously performed epigenome-wide association studies (**Supplementary Table 7**).

Accordingly, for each of these five models, 5-repeated 5-fold cross-validation was applied to find the optimal hyperparameter values that yield the maximal area under the receiver operating characteristic curve (AUROC) (for discrete risk factors) or minimal MAE (for continuous risk factors) (the searched hyperparameter space is shown in **Supplementary Table 6**). From the five models per risk factor, the model that achieved the highest AUROC or R^2^ over all folds was considered as the best-performing risk factor model.

Subsequently, the predicted risk scores of each risk factor model (for binary variables this is defined as log(1/1-p), where p is the estimated class probability), referred to as methylation profile scores (MPSs), were used as variables for the construction of a multivariate methylation risk score (MMRS) for the prediction of *‘MCI versus control’* (*i.e.,* MMRS-MCI model) and *‘AD versus control’* (*i.e.,* MMRS-AD model) in the training set of the EMIF-AD cohort. For this, the Kennard-Stone algorithm [56] was first applied to the MPSs to split the data into a training (n = 436) and an independent test set (n = 187) (**Supplementary Table 8**). Accordingly, an ElasticNet (EN), sparse partial least squares-discriminant analysis (sPLS-DA), and Random Forest model with recursive feature elimination (RF-RFE) models were trained by 5-repeated 5-fold cross-validation to find the optimal hyperparameter combination that yields the highest AUROC (the searched hyperparameter space is shown in **Supplementary Table 9**).

### 2.4. Model validation

In this step, besides the estimation of the model’s performance in the independent test set of the EMIF-AD cohort, the validation of our best-performing model includes the survival analysis of cognitive impairments in the PPMI and ADNI longitudinal cohorts.

Based on the risk scores calculated by our best-performing model among the epi-LIBRA, epi-CAIDE and MMRS models, the individuals of the ADNI and PPMI cohorts were categorized into three equally sized risk categories; low- (n = 74 for ADNI, n = 43 for PPMI), intermediate- (n = 75 for ADNI, n = 43 for PPMI), and high-risk (n = 74 for ADNI and n = 43 for PPMI). We accordingly assessed the statistical significance of the difference in time-dependent conversion to cognitive impairments (as defined in *section 2.2.1*.) by comparing the low- and high-risk groups using the *log-rank* test and a Cox regression model (*survival* package (v3.5.5) [57]). Furthermore, a Kaplan-Meier curve was constructed to visualize the probability of cognitive impairments over time for each of the three risk categories. Please note that in the ADNI cohort, not all cognitive measures were available for each individual and, hence, the number of samples per risk category was different per cognitive outcome (**Supplementary Table 2**).

### 2.5. Model interpretation

#### 2.5.1. Variable importance

The importance of the best-performing model’s variables was evaluated using mean absolute SHapley Additive exPlanations (SHAP) values of the test set samples in the EMIF-AD cohort as calculated with the *DALEX* package (v2.4.3) [58]. Notably, to make the SHAP values comparable between models, the values were normalized such that the absolute sum equals one. The scaled mean absolute SHAP values can therefore be interpreted as the average proportional contribution to the predicted score.

#### 2.5.2. GO overrepresentation analysis

The *missMethyl* package (v1.34.0) [59] was used to perform GO overrepresentation analysis on the union of the most important features of the risk factor models that are used for the prediction by the best-performing model. For the ElasticNet models, the most important features are the CpGs with a non-zero coefficient, while for the Random Forest models, the 1000 CpGs with the largest Gini index were considered as the most important variables.

#### 2.5.3. Influence of genetic variation

To test the extent to which the variation in the methylation status of the model’s CpGs can be explained by genetic variation, the *joint and individual variation explained* (JIVE) method (*r.jive* package (v2.4) [60]) was performed on the risk factor model’s most important features and the corresponding methylation quantitative trait loci (mQTLs) (clumped cis- and trans-mQTLs, P value < 1e-5, GoDMC database [19]). The joint variation, as quantified by the JIVE method, of the risk factor model’s CpG β-values and their mQTLs was recorded.

In addition, colocalization analysis (*coloc* package (v5.2.2) [61]) was performed for each of the model’s most important features to assess whether its methylation status and Alzheimer’s disease status share a common genetic variant. For this, the GWAS summary statistics of Alzheimer’s disease from Marioni *et al.* (2018) [45] and the mQTL data (non-clumped cis-mQTLs, P value < 1e-5) from the GoDMC database [19] were used.

### 2.6. Model extension

Finally, we performed training in the EMIF-AD cohort using the 14 MPSs with 12 PGSs (**Supplementary Table 5**) and/or three CSF biomarkers (*i.e.,* Aβ, t-tau, and p-tau z-scores) as additional variables. For this, the same machine learning strategy as described in *section 2.3.2*. was applied.

## 3. RESULTS

### 3.1. Generation and validation of the epi-CAIDE and epi-LIBRA scores (approach 1)

While the CAIDE score could be relatively well predicted by the Random Forest model with a cross-validation R^2^ ≈ 0.47, the LIBRA score was poorly predicted with a maximal cross-validation R^2^ ≈ 0.04 by the Random Forest model (**Supplementary Table 10**).

Interestingly, in the EMIF-AD cohort, the epi-CAIDE score was found to be highly correlated with chronological (R^2^ ≈ 0.45) and epigenetic (R^2^ ≈ 0.55) age, indicating that age is the main driver of the epi-CAIDE score. Nevertheless, with an AUROC ≤ 0.61, both the epi-CAIDE and epi-LIBRA scores as predicted by the best-performing Random Forest models were shown to be poor estimators of both MCI and AD status in the independent test set of the EMIF-AD cohort (Fig. 2 and **Supplementary Table 11**).

**FIGURE 2.**
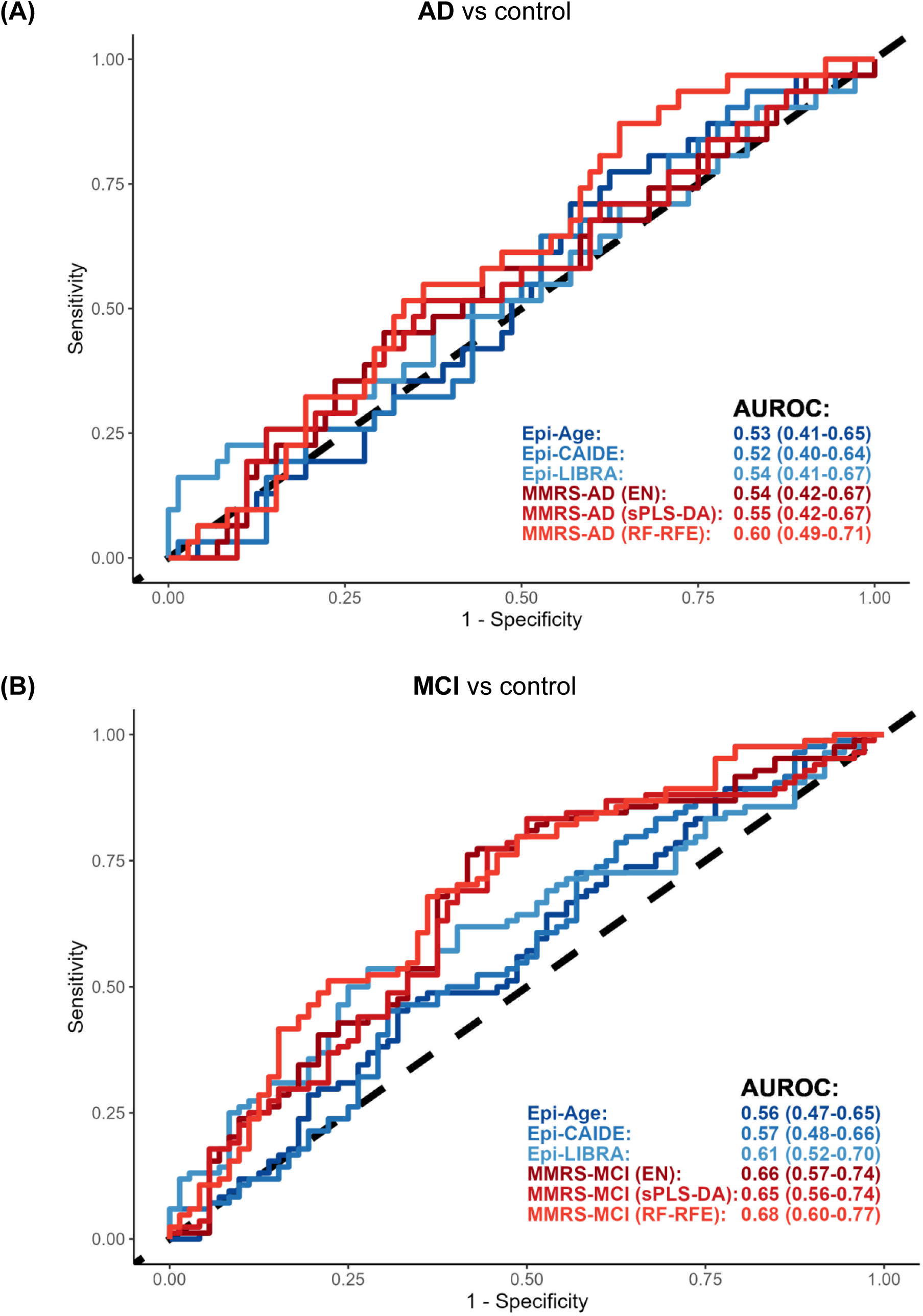
ROC curves of AD and MCI prediction in the independent test set of the EMIF-AD cohort. The MMRS models (red) are trained on the 14 MPSs for the prediction of AD **(A)** and MCI **(B)**. The epi-LIBRA and epi-CAIDE scores (blue) are both predicted by the Random Forest model with correlation-based feature selection method (*i.e.,* the best performing model) from the EXTEND data. The 95% confidence intervals of the AUROC values are indicated between brackets.

### 3.2. Generation and validation of MMRS models (approach 2)

Besides age (R^2^ ≈ 0.92) and sex (AUROC = 1), the best-predicted dementia risk factors by blood-derived DNA methylation data in the EXTEND cohorts include smoking (AUROC ≈ 0.91), type II diabetes (AUROC ≈ 0.89), and heart disease status (AUROC ≈ 0.80) (**Supplementary Tables 12 and 13**). The performance of 9 MPSs could also be validated in the EMIF-AD cohort, demonstrating a mostly lower but statistically significant predictive performance. Just as in the EXTEND cohort, age (R^2^ ≈ 0.87), sex (AUROC = 1), smoking (AUROC ≈ 0.80), and heart disease status (AUROC ≈ 0.67) were found to be the best predicted risk factors in the EMIF-AD cohort (**Supplementary Table 12**).

As shown in Fig. 2 and **Supplementary Table 11**, our MMRS models, which use the 14 MPSs as variables, could not be used to significantly predict AD status in the independent test set of the EMIF-AD cohort (AUROC ≤ 0.60). Nevertheless, our MMRS model (RF-RFE) for MCI was able to significantly predict MCI status with an AUROC of 0.68 (P value ≈ .002, adjusted for age and sex) and significantly improve upon the MCI prediction by epigenetic age (DeLong’s P value ≈ 8.0e-3).

### 3.3. Validation in longitudinal dementia cohorts

To validate our best-performing MMRS model, we performed survival analysis in the ADNI and PPMI longitudinal cohorts. Specifically, for each cohort, we divided the individuals into three equally sized risk categories based on the baseline risk score predicted by the MMRS-MCI (RF-RFE) model and recorded their conversion to cognitive impairments over time.

The Kaplan-Meier curve in Fig. 3A shows that, in the PPMI cohort, there is more conversion to cognitive impairments (*i.e.,* MCI or dementia) within the high-risk group as compared to the low-risk category (log-rank test P value ≈ 6.0e-4, AUROC ≈ 0.67, hazard ratio (HR) [95% confidence interval (CI)] ≈ 3.72 [1.66-8.33]), suggesting that the baseline risk score predicted by the MMRS-MCI model in this Parkinson’s disease cohort is predictive of the onset of cognitive impairments. Furthermore, in the ADNI cohort, we also see more conversion to cognitive impairments as measured by several cognitive tests, reaching statistical significance for *RAVLT - learning* (log-rank test P value ≈ .02, AUROC ≈ 0.62, HR [95% CI] ≈ 2.11 [1.08-4.11]) and *TRABSCOR* (log-rank test P value ≈ .04, AUROC ≈ 0.60, HR [95% CI] ≈ 1.94 [1.01-3.73]) (Fig. 3B, **Supplementary Table 14**, and **Supplementary** Fig 1.).

**FIGURE 3.**
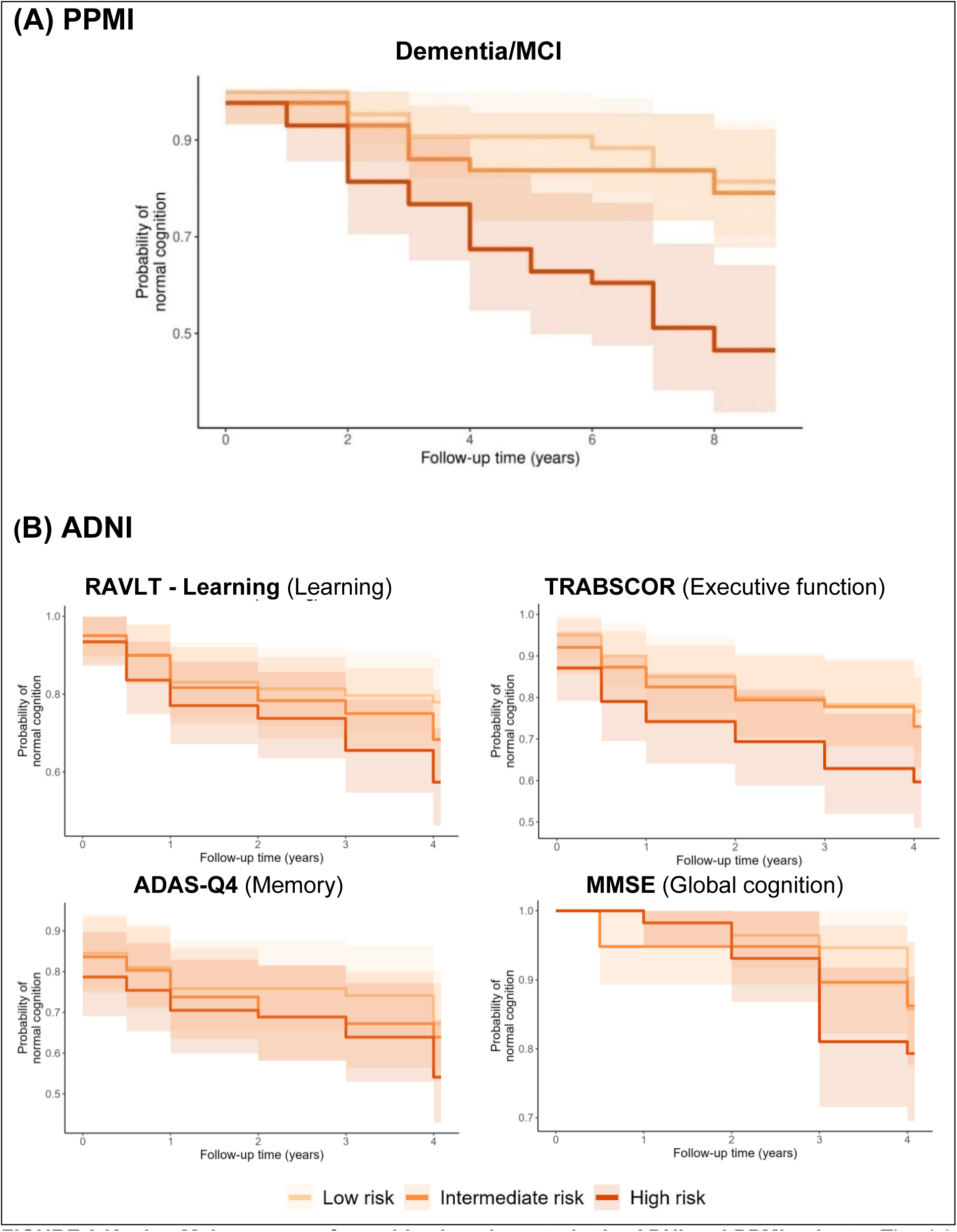
Kaplan-Meier curves of cognitive impairments in the ADNI and PPMI cohorts. The risk categories were defined based on the baseline score predicted by the MMRS-MCI (RF-RFE) model. The shaded area around the line indicates the 95% confidence interval.

### 3.4. Model interpretation

The best performing MMRS-MCI (RF-RFE) model uses only 10 of the 14 MPSs for its prediction. Furthermore, the mean absolute SHAP values indicate that the MMRS is mainly driven by the depression, HDL cholesterol, physical inactivity, and low education MPSs (Fig. 4). The distribution of the MPSs among the diagnostic groups is shown in **Supplementary** Fig 2.

**FIGURE 4.**
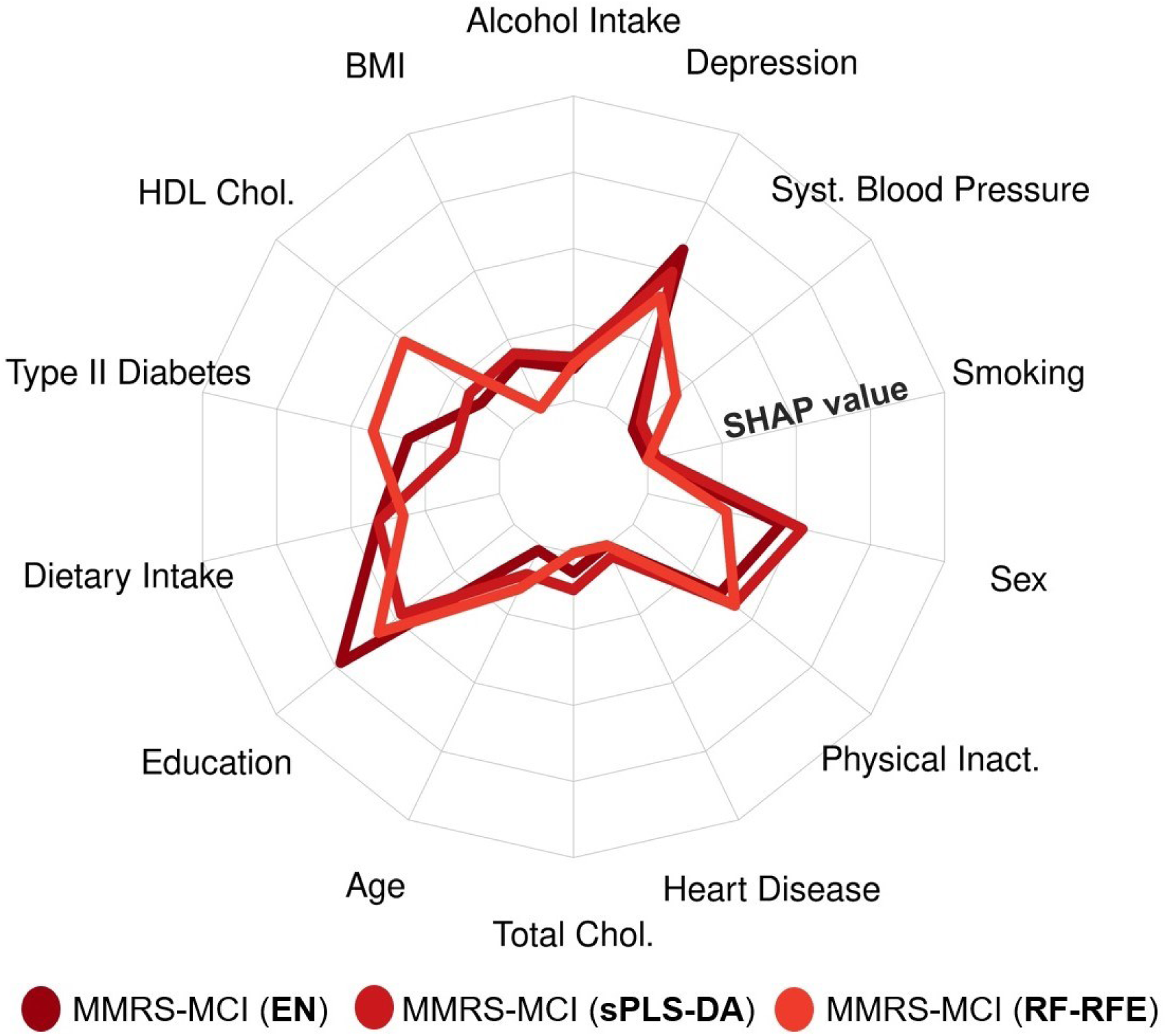
Radar chart of the scaled mean absolute SHAP values. The scaled mean absolute SHAP values indicate the variable importance of the 14 MPSs in the three MMRS-MCI models.

Interestingly, *‘AMPA glutamate receptor clustering (GO:0097113)’* is the most overrepresented GO term by the union of the CpGs used in these 10 risk factor models (7571 CpGs, unadjusted P value ≈ 4.2e-4) (**Supplementary Table 15**). Notably, the model’s CpGs that are associated with this GO term are located in the untranslated region (UTR) or gene body of the *APOE*, *DLG*, *NLGN1*, *SHANK3*, *SHISA6*, *SHISA7*, *SLC7A11*, and *SSH1* genes (**Supplementary** Fig. 3). However, JIVE indicates that there is only minimal joint information captured by the model’s CpGs and their mQTLs. Particularly, for each risk factor model, less than three percent of the variance in DNA methylation data is explained by genetic variation (**Supplementary** Fig. 4). Finally, the colocalization analysis shows that only a few CpGs are likely to share a genetic variant with AD status, including *cg19514613* in the 5’UTR of the *APOE* gene (posterior probability > 0.99) and *cg08374890* located approximately 2 kb upstream of the *BCKDK* gene (posterior probability ≈ 0.94) (**Supplementary** Fig. 5). Together, these results indicate that our best performing model encompasses information unique to DNA methylation data with only a limited genetic influence.

### 3.5. Model extension

Although most PGSs are significantly associated with their corresponding risk factor in the EXTEND and EMIF-AD cohorts, their predictive power is relatively low with an R^2^ ≤ 0.1 and AUROC ≤ 0.67 (**Supplementary Table 16**). In addition, most of the PGSs and their corresponding MPSs have only low correlations (*i.e.,* Pearson correlation coefficient < 0.1) (**Supplementary** Fig. 6).

Furthermore, with an AUROC of 0.69, the addition of 12 PGSs to the 14 MPSs as additional features did not yield a significant improvement in MCI predictive performance above the prediction by MPSs only (Fig. 5). However, the addition of CSF biomarkers to the 14 MPSs could increase the MCI predictive performance to an AUROC of 0.88.

**FIGURE 5.**
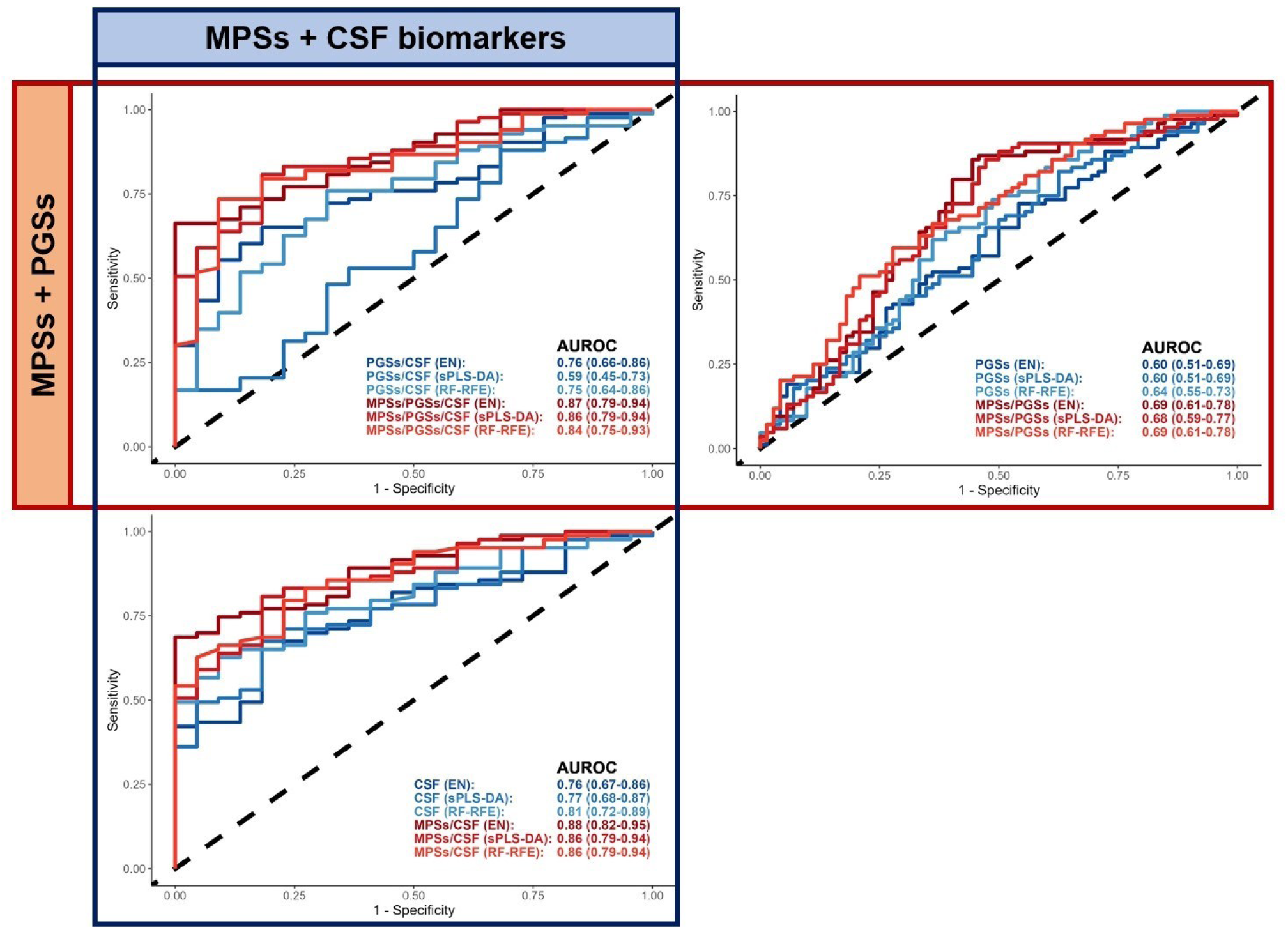
ROC curves of MCI prediction. MCI status was predicted in the independent test set of the EMIF-AD cohort using the MPSs, PGSs, and/or CSF biomarkers as variables.

## 4. DISCUSSION

In this study, we used whole-blood-derived DNA methylation data quantified in a general population of individuals at midlife stage (EXTEND) to create a molecular score as a proxy for CAIDE and LIBRA dementia risk scores (referred to as the epi-CAIDE and epi-LIBRA models, respectively). Alternatively, we generated individual MPSs for 14 modifiable and non-modifiable risk factors contributing to the formation of CAIDE and LIBRA scores. Next, along with epi-CAIDE and epi-LIBRA scores, we trained *MMRS* models using the 14 MPSs in the EMIF-AD training set to predict the AD and MCI status. Subsequently the performance of each model was evaluated in an independent test set. Although our epi-CAIDE and epi-LIBRA models were shown to perform poorly for both AD and MCI prediction in the EMIF-AD cohort, the MMRS model was demonstrated to significantly predict MCI status with an AUROC of 0.68. The potential of this model was further demonstrated by its usefulness for prospectively predicting the onset of cognitive impairments in two independent cohorts of AD (ADNI) and PD (PPMI) as well as improving upon its cross-sectional prediction in combination with established genetic and CSF biomarkers in the EMIF-AD cohort.

The poor performance of our epi-CAIDE and epi-LIBRA models might be explained by the fact that the risk factor’s weights in the CAIDE and LIBRA scores do not consider the extent to which the risk factors can be estimated by DNA methylation data, whereas an ideal epigenetic model would give more weight to the factors best predicted by the DNA methylation data. In addition, some people may have the same CAIDE or LIBRA score but different contributing risk factors. For example, one can have a high CAIDE score because of having a high BMI and low physical activity, while another person may have the same CAIDE score but because of having a high age. These difference in contribution to the total scores may not be effectively captured by our molecular-based epi-CAIDE and epi-LIBRA models, possibly explaining their poor performance.

Similarly, our MMRS model – a model generated from 14 dementia risk factor models – could neither be used to significantly predict AD status in the independent test set of the EMIF-AD cohort (Fig. 2). This poor performance might be explained by the fact that many of the risk factors included in our analysis are cardiovascular-related risk factors (*e.g.,* BMI, systolic blood pressure, heart disease, type II diabetes, physical activity, diet, smoking, as well as HDL and total cholesterol) which might be more predictive of MCI and non-AD types of dementia, like vascular dementia, than AD.

Nevertheless, the MMRS model for MCI, was demonstrated to be able to significantly predict MCI status with an AUROC of up to 0.68 (Fig. 2) as well as to prospectively predict cognitive impairments in two external cohorts (Fig. 3). Noteworthily, despite not being trained in the PPMI cohort, the performance of our best performing MMRS model in prospectively predicting cognitive impairments in this cohort (AUROC ≈ 0.67) was even better than the previously reported prediction by genetic and epigenetic variables (AUROC ≈ 0.64) [30].

Furthermore, compared to using only CSF biomarkers as variables, the combination of the CSF biomarkers and the 14 MPSs resulted in a consistent improvement in the prediction of MCI status (Fig. 5). This combination outperforms previously reported MRI-based classification of MCI in late-life depression [62] and at midlife [63] which have been shown to yield an AUROC of 0.77 and 0.85, respectively. However, the addition of the PGSs as additional features to the 14 MPSs, did not lead to a significant improvement in predictive performance. This is, however, not unforeseen given the relatively poor performance of the PGSs in predicting their corresponding risk factors (**Supplementary Table 16**) and MCI status (Fig. 5).

Although the addition of PGSs yielded only limited improvement in MCI predictive performance, matching pairs of PGSs and MPSs were found to have only a relatively low correlation (**Supplementary** Fig. 6), meaning that the overlapping information between matching pairs of MPSs and PGSs is very limited. Furthermore, JIVE and the colocalization analysis also suggested relatively little influence of genetic variation on the model’s prediction (**Supplementary** Fig. 4-5). Finally, the fact that the addition of MPSs to the PGSs and/or CSF biomarkers consistently improves the model’s performance (Fig. 5), indicates that the MPSs provide unique information on top of the already established genetic and CSF biomarkers.

The ‘*AMPA glutamate receptor clustering (GO:0097113)’* term was shown to be the most significantly overrepresented GO term by the union of the CpGs included in our best-performing MMRS model (**Supplementary Table 15**). Interestingly, AMPA glutamate receptors are known to play an important role in synaptic transmission, and Alzheimer’s disease pathology has previously been shown to be associated with an enhanced removal of these receptors from the post-synaptic membrane [64]. Although the methylation changes in these processes were measured in the blood, they may resemble the alterations that occur in an AD-affected brain. Specifically, the model’s CpGs associated with this GO term are located in known dementia-related genes, including *APOE* [65], *DLG1* [66], *NLGN1* [67], *SHANK3* [68], *SHISA6* [69], *SHISA7* [70], and *SSH1* [71]. Hence, this AMPA receptor-associated process might be one way by which the model is able to estimate a person’s dementia risk.

Besides the validation of our blood-derived DNA methylation-based model in multiple independent cohorts, a strength of the present study is the use of 14 MPSs as variables for dementia risk prediction. Particularly, our approach utilizes the large-scale nature of a general population cohort to reduce the dimensionality of the DNA methylation data into 14 interpretable latent features (*i.e.,* the 14 MPSs), allowing for the construction of a robust and replicable model and overcoming the lack of replication of CpG-level models as described previously [20]. In addition, the MPSs have a clear biological meaning which is in contrast to the more difficult-to-interpret latent features generated by other commonly used dimensionality reduction methods such as (s)PLS, PCA, and autoencoders. This way, our model can provide direct information about which risk factors contribute to an (elevated) dementia risk. As the DNA methylation profile has previously been shown to be modifiable through lifestyle changes (*e.g.,* smoking cessation [72] and weight restoration [73], alcohol withdrawal [74], and physical activity [8]), the information provided by our model possibly allows for targeted intervention strategies, aiming at maximally reducing the patient-specific risk scores. However, to which extent and how these MPSs can be best modified by, for example, lifestyle interventions could be investigated in future studies.

Furthermore, it should be noted that, although MCI is a well-established dementia risk factor, it is not a perfect predictor of the future onset of dementia. Specifically, a significant proportion of MCI individuals revert to a cognitively healthy status [75]. So, instead of using the cross-sectional MCI status as the dependent variable, modeling the prospective cognitive outcome (*e.g.,* the trajectory of cognitive decline) may result in a better model for the prediction of the future onset of cognitive impairments. The lack of large-scale prospective data, however, makes this approach currently impossible, and it is up to future initiatives to collect such data for more sophisticated analyses.

Nevertheless, our established MMRS model may act as a starting point for future studies, aiming at further improving the model’s predictive performance by testing novel feature selection and machine learning methods, incorporating more omics layers, as well as performing model training on larger (prospective) datasets. In the end, this might bring us closer to the establishment of a reliable blood-based model for the early identification of people at risk of developing dementia, an essential requirement for successful intervention strategies.

## Supporting information

Koetsier et al 2023_Manuscript.pdf

## Data Availability

All data produced in the present study are available upon reasonable request to the authors

https://zenodo.org/record/8306113

## ACKNOWLEDGMENTS

This work was supported through a ZonMw Memorabel/Alzheimer Nederland Grant (733050516) to E.P. The current study was conducted as part of the EMIF-AD MBD project, which has received support from the Innovative Medicines Initiative Joint Undertaking under EMIF grant agreement number 115372, the resources of which are composed of financial contribution from the European Union’s Seventh Framework Program (FP7/2007–2013) and EFPIA companies’ in kind contribution. The analysis of the DNA methylation data was in part funded by a major project grant from the Alzheimer’s Society UK (AS-PG-14-038) to KL, a project grant from the Medical Research Council (MRC) (MR/N027973/1) to KL as part of the EPI-AD consortium through the Joint Programme—Neurodegenerative Disease Research (JPND) initiative. Part of this work was supported by the EU’s Horizon2020 program as part of the “Lifebrain” consortium project (to LB). RV acknowledges the support by the Stichting Alzheimer Onderzoek (#13007, #11020, #2017-032) and the Flemish Government (VIND IWT 135043). HZ is a Wallenberg Academy Fellow supported by grants from the Swedish Research Council (#2018-02532), the European Research Council (#681712) and Swedish State Support for Clinical Research (#ALFGBG-720931). SJBV received funding from the Innovative Medicines Initiative 2 Joint Undertaking under ROADMAP grant agreement No. 116020 and from ZonMw during the conduct of this study. No conflict of interest exists. Research at VIB-UAntwerp was in part supported by the University of Antwerp Research Fund. The research was supported by ALF clinical grants from Region Västra Götaland to AW and to PK. The authors acknowledge the assistance of Ellen De Roeck, Naomi De Roeck and Hanne Struyfs (UAntwerp) with data collection. The Lausanne study was funded by a grant from the Swiss National Research Foundation (SNF 320030_141179) to JP. We thank Mrs. Tanja Wesse and Mrs. Sanaz Sedghpour Sabet at the Institute of Clinical Molecular Biology, Christian-Albrechts-University of Kiel, Kiel, Germany for technical assistance with the EPIC methylation profiling. The LIGA team acknowledges computational support from the OMICS compute cluster at the University of Lübeck. We thank all participants and teams who contributed data to PPMI. PPMI, a public-private partnership, is funded by the Michael J. Fox Foundation for Parkinson’s Research and funding partners including 4D Pharma, AbbVie, AcureX Therapeutics, Allergan, Amathus Therapeutics, Aligning Science Across Parkinson’s (ASAP), Avid Radiopharmaceuticals, Bial Biotech, Biogen, BioLegend, Bristol Myers Squibb, Calico Life Sciences LLC, Celgene Corporation, DaCapo Brainscience, Denali Therapeutics, The Edmond J. Safra Foundation, Eli Lilly and Company, GE Healthcare, GlaxoSmithKline, Golub Capital, Handl Therapeutics, Insitro, Janssen Pharmaceuticals, Lundbeck, Merck & Co., Meso Scale Diagnostics LLC, Neurocrine Biosciences, Pfizer, Piramal Imaging, Prevail Therapeutics, F. Hoffmann-La Roche and its affiliated company Genentech, Sanofi Genzyme, Servier, Takeda Pharmaceutical Company, Teva Neuroscience, UCB, Vanqua Bio, Verily Life Sciences, Voyager Therapeutics and Yumanity Therapeutics.

Data collection and sharing for this project was funded by the Alzheimer’s Disease Neuroimaging Initiative (ADNI) (National Institutes of Health Grant U01 AG024904) and DOD ADNI (Department of Defense award number W81XWH-12-2-0012). ADNI is funded by the National Institute on Aging, the National Institute of Biomedical Imaging and Bioengineering, and through generous contributions from the following: AbbVie, Alzheimer’s Association; Alzheimer’s Drug Discovery Foundation; Araclon Biotech; BioClinica, Inc.; Biogen; Bristol-Myers Squibb Company; CereSpir, Inc.; Cogstate; Eisai Inc.; Elan Pharmaceuticals, Inc.; Eli Lilly and Company; EuroImmun; F. Hoffmann-La Roche Ltd and its affiliated company Genentech, Inc.; Fujirebio; GE Healthcare; IXICO Ltd.;Janssen Alzheimer Immunotherapy Research & Development, LLC.; Johnson & Johnson Pharmaceutical Research & Development LLC.; Lumosity; Lundbeck; Merck & Co., Inc.;Meso Scale Diagnostics, LLC.; NeuroRx Research; Neurotrack Technologies; Novartis Pharmaceuticals Corporation; Pfizer Inc.; Piramal Imaging; Servier; Takeda Pharmaceutical Company; and Transition Therapeutics. The Canadian Institutes of Health Research is providing funds to support ADNI clinical sites in Canada. Private sector contributions are facilitated by the Foundation for the National Institutes of Health (www.fnih.org). The grantee organization is the Northern California Institute for Research and Education, and the study is coordinated by the Alzheimer’s Therapeutic Research Institute at the University of Southern California. ADNI data are disseminated by the Laboratory for Neuro Imaging at the University of Southern California.

## DECLARATION OF INTEREST

Declarations of interest: none.

## CODE AVAILABILITY STATEMENT

All codes and an R Shiny application to run the models are available at https://github.com/jarnokoetsier/DementiaRiskPrediction

## Abbreviations

Aβ: amyloid-β
AD: Alzheimer’s disease
ADAS: Alzheimer’s disease assessment scale
AUROC: area under the receiver operating curve
CI: confidence interval
CSF: cerebral spinal fluid
EN: ElasticNet
GO: gene ontology
HR: hazard ratio
HWE: Hardy-Weinberg equilibrium
LDELTOTAL: Wechsler logical memory delay
MAE: mean absolute error
MAF: minor allele frequency
MCI: mild cognitive impairments
mQTL: methylation quantitative trait loci
MMRS: multivariate methylation risk score
MMSE: Mini Mental State Examination
MPS: methylation profile score
PCA: principal component analysis
PD: Parkinson’s disease
PGS: polygenic (risk) score
p-tau: phosphorylated tau
QC: quality control
RAVLT: Rey’s auditory verbal learning test
RF-RFE: random forest with recursive feature elimination
SHAP: Shapley additive explanations
SNP: single nucleotide polymorphism
sPLS-DA: sparse partial least squares discriminant analysis
TRABSCOR: trail making test part B time
t-tau: total tau
UTR: untranslated region.

## Notes

### Competing Interest Statement

The authors have declared no competing interest.

### Author Declarations

https://exetercrfnihr.org/about/exeter-10000-prb/ https://ida.loni.usc.edu/http://www.emif.eu/emif-ad-2/

## REFERENCES

[1] Nichols E, Steinmetz JD, Vollset SE, Fukutaki K, Chalek J, Abd-Allah F, et al. Estimation of the global prevalence of dementia in 2019 and forecasted prevalence in 2050: an analysis for the Global Burden of Disease Study 2019. The Lancet Public Health. 2022;7:e105–e25.

[2] Tisher A, Salardini A. A comprehensive update on treatment of dementia. Seminars in neurology: Thieme Medical Publishers; 2019. p. 167–78.

[3] McGrowder DA, Miller F, Vaz K, Nwokocha C, Wilson-Clarke C, Anderson-Cross M, et al. Cerebrospinal fluid biomarkers of Alzheimer’s disease: current evidence and future perspectives. Brain Sciences. 2021;11:215.

[4] Jo T, Nho K, Saykin AJ. Deep learning in Alzheimer’s disease: diagnostic classification and prognostic prediction using neuroimaging data. Frontiers in aging neuroscience. 2019;11:220.

[5] Kim JH, Chang IB, Kim YH, Min CY, Yoo DM, Choi HG. Association Between Various Types or Statuses of Smoking and Subjective Cognitive Decline Based on a Community Health Survey of Korean Adults. Frontiers in neurology. 2022:901.

[6] Schaefer SM, Kaiser A, Behrendt I, Eichner G, Fasshauer M. Association of Alcohol Types, Coffee, and Tea Intake with Risk of Dementia: Prospective Cohort Study of UK Biobank Participants. Brain Sciences. 2022;12:360.

[7] Malik R, Georgakis MK, Neitzel J, Rannikmäe K, Ewers M, Seshadri S, et al. Midlife vascular risk factors and risk of incident dementia: Longitudinal cohort and Mendelian randomization analyses in the UK Biobank. Alzheimer’s & Dementia. 2021;17:1422–31.

[8] Iso-Markku P, Waller K, Vuoksimaa E, Heikkilä K, Rinne J, Kaprio J, et al. Midlife physical activity and cognition later in life: a prospective twin study. Journal of Alzheimer’s Disease. 2016;54:1303–17.

[9] Xu W, Tan L, Wang H-F, Tan M-S, Tan L, Li J-Q, et al. Education and risk of dementia: dose-response meta-analysis of prospective cohort studies. Molecular neurobiology. 2016;53:3113–23.

[10] Shannon OM, Ranson JM, Gregory S, Macpherson H, Milte C, Lentjes M, et al. Mediterranean diet adherence is associated with lower dementia risk, independent of genetic predisposition: findings from the UK Biobank prospective cohort study. BMC medicine. 2023;21:1–13.

[11] Kivipelto M, Ngandu T, Laatikainen T, Winblad B, Soininen H, Tuomilehto J. Risk score for the prediction of dementia risk in 20 years among middle aged people: a longitudinal, population-based study. The Lancet Neurology. 2006;5:735–41.

[12] Schiepers OJ, Köhler S, Deckers K, Irving K, O’Donnell CA, van den Akker M, et al. Lifestyle for Brain Health (LIBRA): a new model for dementia prevention. International journal of geriatric psychiatry. 2018;33:167–75.

[13] Hansson O, Edelmayer RM, Boxer AL, Carrillo MC, Mielke MM, Rabinovici GD, et al. The Alzheimer’s Association appropriate use recommendations for blood biomarkers in Alzheimer’s disease. Alzheimer’s & Dementia. 2022;18:2669–86.

[14] Fransquet PD, Lacaze P, Saffery R, McNeil J, Woods R, Ryan J. Blood DNA methylation as a potential biomarker of dementia: a systematic review. Alzheimer’s & Dementia. 2018;14:81–103.

[15] Lim U, Song MA. Dietary and lifestyle factors of DNA methylation. Methods in molecular biology (Clifton, NJ). 2012;863:359–76.

[16] Joehanes R, Just AC, Marioni RE, Pilling LC, Reynolds LM, Mandaviya PR, et al. Epigenetic signatures of cigarette smoking. Circulation: cardiovascular genetics. 2016;9:436–47.

[17] Samblas M, Milagro FI, Martínez A. DNA methylation markers in obesity, metabolic syndrome, and weight loss. Epigenetics. 2019;14:421–44.

[18] Gonzalez-Jaramillo V, Portilla-Fernandez E, Glisic M, Voortman T, Bramer W, Chowdhury R, et al. The role of DNA methylation and histone modifications in blood pressure: a systematic review. Journal of human hypertension. 2019;33:703–15.

[19] Min JL, Hemani G, Hannon E, Dekkers KF, Castillo-Fernandez J, Luijk R, et al. Genomic and phenotypic insights from an atlas of genetic effects on DNA methylation. Nature genetics. 2021;53:1311–21.

[20] Josefsson M, Landfors M, Kauppi K, Porter T, Milicic L, Laws S, et al. Sixteen-Year Longitudinal Evaluation of Blood-Based DNA Methylation Biomarkers for Early Prediction of Alzheimer’s Disease. Journal of Alzheimer’s Disease: JAD. 2023.

[21] Hattersley A. The Exeter 10,000 (EXTEND) project. In: Facility NECR, editor.2020.

[22] Bos I, Vos S, Vandenberghe R, Scheltens P, Engelborghs S, Frisoni G, et al. The EMIF-AD Multimodal Biomarker Discovery study: design, methods and cohort characteristics. Alzheimer’s research & therapy. 2018;10:1–9.

[23] Marek K, Chowdhury S, Siderowf A, Lasch S, Coffey CS, Caspell-Garcia C, et al. The Parkinson’s progression markers initiative (PPMI)–establishing a PD biomarker cohort. Annals of clinical and translational neurology. 2018;5:1460–77.

[24] Petersen RC, Aisen PS, Beckett LA, Donohue MC, Gamst AC, Harvey DJ, et al. Alzheimer’s disease neuroimaging initiative (ADNI): clinical characterization. Neurology. 2010;74:201–9.

[25] Petersen RC. Mild cognitive impairment as a diagnostic entity. Journal of internal medicine. 2004;256:183–94.

[26] Winblad B, Palmer K, Kivipelto M, Jelic V, Fratiglioni L, Wahlund LO, et al. Mild cognitive impairment–beyond controversies, towards a consensus: report of the International Working Group on Mild Cognitive Impairment. Journal of internal medicine. 2004;256:240–6.

[27] McKhann G, Drachman D, Folstein M, Katzman R, Price D, Stadlan EM. Clinical diagnosis of Alzheimer’s disease: Report of the NINCDS-ADRDA Work Group* under the auspices of Department of Health and Human Services Task Force on Alzheimer’s Disease. Neurology. 1984;34:939-.

[28] Litvan I, Goldman JG, Tröster AI, Schmand BA, Weintraub D, Petersen RC, et al. Diagnostic criteria for mild cognitive impairment in Parkinson’s disease: Movement Disorder Society Task Force guidelines. Movement disorders. 2012;27:349–56.

[29] Emre M, Aarsland D, Brown R, Burn DJ, Duyckaerts C, Mizuno Y, et al. Clinical diagnostic criteria for dementia associated with Parkinson’s disease. Movement disorders: official journal of the Movement Disorder Society. 2007;22:1689–707.

[30] Harvey J, Reijnders RA, Cavill R, Duits A, Köhler S, Eijssen L, et al. Machine learning-based prediction of cognitive outcomes in de novo Parkinson’s disease. npj Parkinson’s Disease. 2022;8:150.

[31] Stephen R, Ngandu T, Liu Y, Peltonen M, Antikainen R, Kemppainen N, et al. Change in CAIDE dementia risk score and neuroimaging biomarkers during a 2-year multidomain lifestyle randomized controlled trial: Results of a post-hoc subgroup analysis. The Journals of Gerontology: Series A. 2021;76:1407–14.

[32] Aryee MJ, Jaffe AE, Corrada-Bravo H, Ladd-Acosta C, Feinberg AP, Hansen KD, et al. Minfi: a flexible and comprehensive Bioconductor package for the analysis of Infinium DNA methylation microarrays. Bioinformatics. 2014;30:1363–9.

[33] Pidsley R, Y Wong CC, Volta M, Lunnon K, Mill J, Schalkwyk LC. A data-driven approach to preprocessing Illumina 450K methylation array data. BMC genomics. 2013;14:1–10.

[34] Liu J, Siegmund KD. An evaluation of processing methods for HumanMethylation450 BeadChip data. BMC genomics. 2016;17:1–11.

[35] Pidsley R, Zotenko E, Peters TJ, Lawrence MG, Risbridger GP, Molloy P, et al. Critical evaluation of the Illumina MethylationEPIC BeadChip microarray for whole-genome DNA methylation profiling. Genome biology. 2016;17:1–17.

[36] McCartney DL, Walker RM, Morris SW, McIntosh AM, Porteous DJ, Evans KL. Identification of polymorphic and off-target probe binding sites on the Illumina Infinium MethylationEPIC BeadChip. Genomics data. 2016;9:22–4.

[37] Josse J, Husson F. missMDA: a package for handling missing values in multivariate data analysis. Journal of statistical software. 2016;70:1–31.

[38] Lena PD, Sala C, Prodi A, Nardini C. Methylation data imputation performances under different representations and missingness patterns. BMC bioinformatics. 2020;21:1–22.

[39] Hong S, Prokopenko D, Dobricic V, Kilpert F, Bos I, Vos SJB, et al. Genome-wide association study of Alzheimer’s disease CSF biomarkers in the EMIF-AD Multimodal Biomarker Discovery dataset. Transl Psychiatry. 2020;10:403.

[40] McCarthy. HRC or 1000G Imputation preparation and checking. 2018.

[41] Das S, Forer L, Schönherr S, Sidore C, Locke AE, Kwong A, et al. Next-generation genotype imputation service and methods. Nature genetics. 2016;48:1284–7.

[42] Zhang Q, Privé F, Vilhjálmsson B, Speed D. Improved genetic prediction of complex traits from individual-level data or summary statistics. Nature communications. 2021;12:4192.

[43] Yengo L, Sidorenko J, Kemper KE, Zheng Z, Wood AR, Weedon MN, et al. Meta-analysis of genome-wide association studies for height and body mass index in∼ 700000 individuals of European ancestry. Hum Mol Genet. 2018;27:3641–9.

[44] Glucose M-Ao, Investigators I-rtC, Consortium GIoAT, Consortium AGENTD, Consortium SATD, Shuldiner AR, et al. Large-scale association analysis provides insights into the genetic architecture and pathophysiology of type 2 diabetes. Nature genetics. 2012;44:981–90.

[45] Marioni RE, Harris SE, Zhang Q, McRae AF, Hagenaars SP, Hill WD, et al. GWAS on family history of Alzheimer’s disease. Translational psychiatry. 2018;8:99.

[46] Willer CJ, Schmidt EM, Sengupta S, Peloso GM, Gustafsson S, Kanoni S, et al. Discovery and refinement of loci associated with lipid levels. Nat Genet. 2013;45:1274–83.

[47] Howard DM, Adams MJ, Clarke T-K, Hafferty JD, Gibson J, Shirali M, et al. Genome-wide meta-analysis of depression identifies 102 independent variants and highlights the importance of the prefrontal brain regions. Nature neuroscience. 2019;22:343–52.

[48] Okbay A, Wu Y, Wang N, Jayashankar H, Bennett M, Nehzati SM, et al. Polygenic prediction of educational attainment within and between families from genome-wide association analyses in 3 million individuals. Nature genetics. 2022;54:437–49.

[49] Nelson CP, Goel A, Butterworth AS, Kanoni S, Webb TR, Marouli E, et al. Association analyses based on false discovery rate implicate new loci for coronary artery disease. Nature genetics. 2017;49:1385–91.

[50] Niarchou M, Byrne EM, Trzaskowski M, Sidorenko J, Kemper KE, McGrath JJ, et al. Genome-wide association study of dietary intake in the UK biobank study and its associations with schizophrenia and other traits. Translational Psychiatry. 2020;10:51.

[51] Abbott L, Bryant S, Churchhouse C, Ganna A, Howrigan D, Palmer D, et al. UK Biobank GWAS round 2. 2018.

[52] Kranzler HR, Zhou H, Kember RL, Vickers Smith R, Justice AC, Damrauer S, et al. Genome-wide association study of alcohol consumption and use disorder in 274,424 individuals from multiple populations. Nature communications. 2019;10:1499.

[53] Wang Z, Emmerich A, Pillon NJ, Moore T, Hemerich D, Cornelis MC, et al. Genome-wide association analyses of physical activity and sedentary behavior provide insights into underlying mechanisms and roles in disease prevention. Nature genetics. 2022;54:1332–44.

[54] Hillary RF, Marioni RE. MethylDetectR: a software for methylation-based health profiling. Wellcome Open Res. 2020;5:283.

[55] Zhang Q, Vallerga CL, Walker RM, Lin T, Henders AK, Montgomery GW, et al. Improved precision of epigenetic clock estimates across tissues and its implication for biological ageing. Genome medicine. 2019;11:1–11.

[56] Kennard RW, Stone LA. Computer aided design of experiments. Technometrics. 1969;11:137–48.

[57] Therneau TM, Lumley T. Package ‘survival’. R Top Doc. 2015;128:28–33.

[58] Biecek P. DALEX: Explainers for complex predictive models in R. The Journal of Machine Learning Research. 2018;19:3245–9.

[59] Phipson B, Maksimovic J, Oshlack A. missMethyl: an R package for analyzing data from Illumina’s HumanMethylation450 platform. Bioinformatics. 2015;32:286–8.

[60] O’Connell MJ, Lock EF. R. JIVE for exploration of multi-source molecular data. Bioinformatics. 2016;32:2877–9.

[61] Rasooly D, Peloso GM, Giambartolomei C. Bayesian Genetic Colocalization Test of Two Traits Using coloc. Current Protocols. 2022;2:e627.

[62] Lebedeva AK, Westman E, Borza T, Beyer MK, Engedal K, Aarsland D, et al. MRI-based classification models in prediction of mild cognitive impairment and dementia in late-life depression. Frontiers in aging neuroscience. 2017:13.

[63] Ota K, Oishi N, Ito K, Fukuyama H, Group S-JS. A comparison of three brain atlases for MCI prediction. Journal of neuroscience methods. 2014;221:139–50.

[64] Babaei P. NMDA and AMPA receptors dysregulation in Alzheimer’s disease. European Journal of Pharmacology. 2021;908:174310.

[65] Martens YA, Zhao N, Liu CC, Kanekiyo T, Yang AJ, Goate AM, et al. ApoE Cascade Hypothesis in the pathogenesis of Alzheimer’s disease and related dementias. Neuron. 2022;110:1304–17.

[66] Taskesen E, Mishra A, van der Sluis S, Ferrari R, Veldink JH, van Es MA, et al. Susceptible genes and disease mechanisms identified in frontotemporal dementia and frontotemporal dementia with Amyotrophic Lateral Sclerosis by DNA-methylation and GWAS. Sci Rep. 2017;7:8899.

[67] Arias-Aragón F, Tristán-Clavijo E, Martínez-Gallego I, Robles-Lanuza E, Coatl-Cuaya H, Martín-Cuevas C, et al. A Neuroligin-1 mutation associated with Alzheimer’s disease produces memory and age-dependent impairments in hippocampal plasticity. iScience. 2023;26:106868.

[68] Landry O, François A, Oye Mintsa Mi-Mba MF, Traversy MT, Tremblay C, Emond V, et al. Postsynaptic Protein Shank3a Deficiency Synergizes with Alzheimer’s Disease Neuropathology to Impair Cognitive Performance in the 3xTg-AD Murine Model. The Journal of neuroscience : the official journal of the Society for Neuroscience. 2023;43:4941–54.

[69] Ramos J, Caywood LJ, Prough MB, Clouse JE, Herington SD, Slifer SH, et al. Genetic variants in the SHISA6 gene are associated with delayed cognitive impairment in two family datasets. Alzheimer’s & dementia : the journal of the Alzheimer’s Association. 2023;19:611–20.

[70] Sabaie H, Talebi M, Gharesouarn J, Asadi MR, Jalaiei A, Arsang-Jang S, et al. Identification and Analysis of BCAS4/hsa-miR-185-5p/SHISA7 Competing Endogenous RNA Axis in Late-Onset Alzheimer’s Disease Using Bioinformatic and Experimental Approaches. Front Aging Neurosci. 2022;14:812169.

[71] Cazzaro S, Woo JA, Wang X, Liu T, Rego S, Kee TR, et al. Slingshot homolog-1-mediated Nrf2 sequestration tips the balance from neuroprotection to neurodegeneration in Alzheimer’s disease. Proc Natl Acad Sci U S A. 2023;120:e2217128120.

[72] Tsaprouni LG, Yang T-P, Bell J, Dick KJ, Kanoni S, Nisbet J, et al. Cigarette smoking reduces DNA methylation levels at multiple genomic loci but the effect is partially reversible upon cessation. Epigenetics. 2014;9:1382–96.

[73] Steiger H, Booij L, Kahan E, McGregor K, Thaler L, Fletcher E, et al. A longitudinal, epigenome-wide study of DNA methylation in anorexia nervosa: results in actively ill, partially weight-restored, long-term remitted and non-eating-disordered women. Journal of Psychiatry and Neuroscience. 2019;44:205–13.

[74] Witt SH, Frank J, Frischknecht U, Treutlein J, Streit F, Foo JC, et al. Acute alcohol withdrawal and recovery in men lead to profound changes in DNA methylation profiles: a longitudinal clinical study. Addiction. 2020;115:2034–44.

[75] Jongsiriyanyong S, Limpawattana P. Mild Cognitive Impairment in Clinical Practice: A Review Article. American journal of Alzheimer’s disease and other dementias. 2018;33:500–7.

