## Supplementary material for "Blood-based multivariate methylation risk score for cognitive impairment and dementia": Koetsier et al 2023_Manuscript.pdf

- SUPPLEMENTARY MATERIALS -

Content

### Supplementary Tables

| **Cohort** | **Description** | **Training/validation** | **N** | **Sex (M/F)** | **Age (mean ± SD)** |
| --- | --- | --- | --- | --- | --- |
| EXTEND | General population cohort | Model training of MPS, epi-CAIDE, and epi-LIBRA models. | 1076 | 529/547 | 57.1 ± 9.3 |
| EMIF-AD | Alzheimer’s disease cohort | Model training (in training set) of the MMRS models and model validation (in test set) of the MMRS, epi-CAIDE, and epi-LIBRA models. | 662 | 322/340 | 65.8 ± 6.4 |
| PPMI | Parkinson’s disease cohort | Model validation (survival analysis) of the best performing MMRS model. | 129 | 36/93 | 61.2 ± 7.3 |
| ADNI | Alzheimer’s disease cohort | Model validation (survival analysis) of the best performing MMRS model. | 223 | 118/105 | 68.2 ± 4.5 |

**SUPPLEM** **ENTARY TABLE 1 Overview of the cohort characteristics.**

**SUPPLEM** **ENTARY TABLE 2 Cognitive outcomes used for the time analysis in the ADNI cohort.**

| **Cognitive test** | **Description** | **Cognitive domain** | **Cognitive impairment** | **N *** |
| --- | --- | --- | --- | --- |
| **ADAS11** | *Alzheimer’s Disease Assessment Scale; a measure of cognitive dysfunction based on 11 questions*  *.* | Global | > Mean + 2 SD | 62/62/63 |
| **ADAS13** | *Alzheimer’s Disease Assessment Scale; a measure of cognitive dysfunction based on 13 questions.* | Global | > Mean + 2 SD | 62/60/62 |
| **ADASQ4** | *Alzheimer’s Disease Assessment Scale Question 4; delayed word recall.* | **Memory** & Learning | > Mean + 2 SD | 58/61/61 |
| **RAVLT - Learning** | *Rey’s Auditory Verbal Learning Test; learning ability.* | Memory & **Learning** | < Mean - 2 SD | 59/60/61 |
| **RAVLT - Forgetting** | *Rey’s Auditory Verbal Learning Test; delayed memory.* | **Memory** & Learning | > Mean + 2 SD | 55/58/55 |
| **RAVLT - Percent forgetting** | *Rey’s Auditory Verbal Learning Test; delayed memory.* | **Memory** & Learning | > Mean + 2 SD | 61/61/66 |
| **RAVLT - Immediate recall** | *Rey’s Auditory Verbal Learning Test; immediate recall.* | **Memory** & Learning | < Mean - 2 SD | 59/60/61 |
| **LDELTOTAL** | *Wechsler Logical Memory Delay.* | **Memory** & Learning | < Mean - 2 SD | 61/60/64 |
| **TRABSCOR** | *Trail Making Test Part B Time.* | **Executive function** | > Mean + 2 SD | 60/63/62 |
| **MMSE** | *Mini Mental State Examination.* | Global | < 24 | 56/58/58 |

* The number of samples in the low/intermediate/high risk group in the survival analysis.

**SUPPLEMENTARY TABLE 3 Risk factors and their weights used for the calculation of the CAIDE score.**

| **Risk factor** | **Details** | **Groups** | **Score** |
| --- | --- | --- | --- |
| **Age** | *Chronological age.* | Age < 47 years  47 ≤ Age ≤ 53 years  Age > 53 years | **0**  **+3**  **+4** |
| **Sex** | *Sex.* | Female  Male | **0**  **+1** |
| **Education** | *A person is defined to be lowly educated if the individual has none of the following educational achievements:*  *1. College or university degree.*  *2. O level, GCSEs, or equivalent.*  *3. NVQ, HND, HNC, or equivalent.*  *4. A-level, AS-level, or equivalent.*  *5. CSEs or equivalent.*  *6. Other professional qualifications* | Educated  Uneducated | **0**  **+2** |
| **Systolic blood pressure** | *Mean systolic blood pressure as determined by physical examination.* | Systolic blood pressure ≤ 140 mmHg  Systolic blood pressure > 140 mmHg | **0**  **+2** |
| **Body Mass Index (BMI)** | *Body Mass Index (BMI) is defined as a person’s weight (kilograms) divided by the person’s height squared (meters^2^).* | BMI ≤ 30 kg/m^2^  BMI > 30 kg/m^2^ | **0**  **+2** |
| **Serum Total cholesterol** | *Total cholesterol concentration in serum based on biosample assay.* | Serum total cholesterol ≤ 6.5 mmol/L  Serum total cholesterol > 6.5 mmol/L | **0**  **+2** |
| **Physical activity** | *A person is defined to be physical inactive if self-reported to do exercise with increased pulse more than 2.5 hours per week.4* | Physically active  Physically inactive | **0**  **+1** |

**SUPPLEMENTARY TABLE 4 Risk factors and their weights used for the calculation of the LIBRA score.**

| **Risk factor** | **Details** | **Groups** | **Score** |
| --- | --- | --- | --- |
| **Healthy diet** | *Healthy diet is defined as 3 portions fruit and/or 3 portions vegetables per day.* | Healthy diet  Unhealthy diet | **-1.7**  **0** |
| **Physical inactivity** | *A person is defined to be physical inactive if self-reported to do exercise with increased pulse more than 2.5 hours per week.* | Physically active  Physically inactive | **0**  **+1.1** |
| **Smoking** | *Self-reported smoking status.* | No smoker  Current smoker | **0**  **+1.5** |
| **Alcohol intake** | *High alcohol intake is defined by one of the following criteria:*  *1. Once a month, more than 10 alcoholic drinks per day.*  *2. 2-4 a month, 5 or more alcoholic drinks per day*  *3. 2-3 a week, 5 or more alcoholic drinks per day*  *4. 4 or more a week, 3 or more alcoholic drinks per day* | L-M alcohol intake  High alcohol intake | **-1**  **0** |
| **Obesity** | *Body Mass Index (BMI) is defined as a person’s weight (kilograms) divided by the person’s height squared (meters^2^).* | BMI < 30 kg/m^2^  BMI ≥ 30 kg/m^2^ | **0**  **+1.6** |
| **Depression** | *Self-reported depression status* | Non-depressed  Depressed | **0**  **+2.1** |
| **Type II Diabetes** | *Self-reported type II diabetes status* | No type II diabetes  Type II diabetes | **0**  **+1.3** |
| **Hypertension** | *An individual’s hypertension status is based on the measured systolic and diastolic blood pressured.* | Systolic blood pressure ≤ 140 mmHg & Diastolic blood pressure ≤ 90 mmHg  Systolic blood pressure > 140 mmHg or  Diastolic blood pressure > 90 mmHg | **0**  **+1.6** |
| **High HDL Cholesterol** | *High-Density Lipoprotein (HDL) concentration in serum based on biosample assay.* | HDL ≤ 2.2 mmol/L  HDL > 2.2 mmol/L | **0**  **+1.4** |
| **Heart Disease** | *Self-reported heart disease status* | No heart disease  Heart disease | **0**  **+1** |
| **Kidney Disease** | *Self-reported chronic kidney disease status* | No kidney disease  Kidney disease | **0**  **+1.1** |

**SUPPLEMENTARY TABLE 5 Traits and corresponding publication for which the polygenic (risk) scores (PGSs) were calculated.**

| Trait | Year | Author | DOI/URL |
| --- | --- | --- | --- |
| *BMI (Body Mass Index)* | 2018 | Yengo *et al.* [1] | <https://doi.org/10.1093/hmg/ddy271> |
| *T2D (Type II Diabetes)* | 2012 | DIAGRAM consortium [2] | <https://doi.org/10.1038%2Fng.2383> |
| *AD (Alzheimer’s Disease)* | 2018 | Marioni *et al.* [3] | <https://doi.org/10.1038/s41398-019-0498-2> |
| *HDL (High-Density Lipoprotein) Cholesterol* | 2013 | GLG consortium [4] | <https://doi.org/10.1038/ng.2797> |
| *TC (Total Cholesterol)* | 2013 | GLG consortium [4] | <https://doi.org/10.1038/ng.2797> |
| *MDD (major depression disorder)* | 2019 | Howard *et al.* [5] | <https://doi.org/10.1038/s41593-018-0326-7> |
| *EA (Educational attainment)* | 2022 | Okbay *et al.* [6] | <https://doi.org/10.1038/s41588-022-01016-z> |
| *CAD (coronary artery disease)* | 2017 | Nelson *et al.* [7] | <https://doi.org/10.1038/ng.3913> |
| *DC2 (Fish- and plant-based diet)* | 2020 | Niarchou *et al.* [8] | <https://doi.org/10.1038/s41398-020-0688-y> |
| *SBP (Systolic blood pressure), automated reading* | 2018 | UK Biobank (UKBB) [9] | <http://www.nealelab.is/uk-biobank> |
| *Alcohol consumption* | 2019 | Kranzler *et al.* [10] | <https://doi.org/10.1038/s41467-019-09480-8> |
| *MVPA (moderate-to-vigorous intensity physical activity during leisure time)* | 2022 | Wang *et al.* [11] | <https://doi.org/10.1038/s41588-022-01165-1> |

**SUPPLEMENTARY TABLE 6** **The search space for the hyperparameter optimization**. The hyperparameters were optimized in a 5-fold 5-repeated cross-validation for the prediction of the continuous (regression) and discrete (classification) risk factors in the EXTEND and EMIF-AD cohort.

| **Prediction** | **Method** | **Parameters** | **Search Space *** |
| --- | --- | --- | --- |
| *Regression* | *ElasticNet-regularized linear regression* | *Alpha* | 0.1 - 1 *(s. l. = 10)* |
|  |  | *Lambda* | 0.01 - 2.5 *(s.l. = 100, exponentially spaced)* |
|  | *Random Forest* | *# Randomly sampled features per tree* | 1000 - 8000 *(s.l. = 8)* |
|  |  | *Minimum node size* | 10 – 60 *(s.l. = 6)* |
|  |  | *Split rule* | Variance |
| *Classification* | *ElasticNet-regularized logistic regression* | *Alpha* | 0.1 - 1 *(s. l. = 10)* |
|  |  | *Lambda* | 0.01 - 2.5 *(s.l. = 100, exponentially spaced)* |
|  | *Random Forest* | *# Randomly sampled features per tree* | 100; 500; 1000; 1500; 2000; 3000; 4000; |
|  |  | *Minimum node size* | 3; 5; 10; 15; 20 |
|  |  | *Split rule* | Gini |

* Note that s.l. is the abbreviation for sequence length and indicates the number of equally spaced values that were tested. For instance, 1 - 5 (s.l. = 5) means that the values 1,2,3,4, and 5 have been tested.

**SUPPLEMENTARY TABLE 7 Number of significant CpGs in the EWASs of the different dementia risk factors.** The significant CpGs were used for model training of the corresponding risk factor (*i.e.,* literature-based feature selection).

| **Risk Factor** | **Publication** | **# CpGs** |
| --- | --- | --- |
| *Body Mass Index (BMI)* | Dhana *et al.* (2018) [12] | 12 |
| *Type II Diabetes* | Fraszczyk *et al.* (2022) [13] | 76 |
| *Alcohol consumption* | Lohoff *et al.* (2022) [14] | 2504 |
| *HDL Cholesterol* | Braun *et al.* (2017) [15] | 56 |
| *Total Cholesterol* | Brain *et al.* (2017) [15] | 4 |
| *Physical Activity* | Fernández-Sanlés *et al.* (2020) [16] | 36 |
| *Coronary Heart Disease (CAD)* | Xia *et al.* (2021) [17] | 52 |
| *Major Depression Disorder (MDD)* | Li *et al.* (2022) [18] | 8 |
| *Educational Attainment (EA)* | Karlsson Linnér *et al.* (2017) [19] | 364 |
| *Dietary Intake*  *(i.e., onions-garlic, nuts-seeds, milk, cream, plant oils, and/or butter intake)* | Hellbach *et al.* (2022) [20] | 126 |
| *(Systolic) Blood Pressure* | Richard *et al.* (2017) [21] | 49 |

| **Model** | **# control samples**  *(training + test set)* | **# MCI samples**  *(training + test set)* | **# AD samples**  *(training + test set)* |
| --- | --- | --- | --- |
| *MPSs* | 148 + 72 | 209 + 84 | 79 + 31 |
| *MPSs + PGSs* | 60 + 22 | 206 + 83 | NA * |
| *MPSs + CSF* | 148 + 72 | 209 + 84 | NA * |
| *MPSs + PGSs + CSF* | 60 + 22 | 206 + 83 | NA * |

**SUPPLEMENTARY TABLE 8** **Number of AD, MCI and control samples in training and test set.**

* Note that AD status was not predicted with PGSs and CSF biomarkers as additional variables.

**SUPPLEMENTARY TABLE 9** **The search space for the hyperparameter optimization.** The hyperparameters were optimized in a 5-fold 5-repeated cross-validation for the prediction of MCI vs control and AD vs control in the EMIF-AD cohort.

| **Method** | **Parameters** | **Search Space *** |
| --- | --- | --- |
| *ElasticNet-regularized logistic regression* | *Alpha* | 0.1 - 1 *(s. l. = 10)* |
|  | *Lambda* | 0.01 - 2.5 *(s.l. = 100, exponentially spaced)* |
| *Sparse Partial Least Squares Discriminant Analysis (sPLS-DA)* | *# Components* | 1 - 10 *(s.l. = 10)* |
|  | *Eta* | 0.1 - 0.9 *(s.l. = 20)* |
|  | *Kappa* | 0.5 |
| *Random Forest – recursive feature elimination †* | *# Randomly sampled features per tree* | 1 - # variables (s.l. = # variables) |
|  | *Minimum node size* | 1 - # variables (s.l. = # variables) |
|  | *Split rule* | Gini |

* Note that s.l. is the abbreviation for sequence length and indicates the number of equally spaced values that were tested. For instance, 1 - 5 (s.l. = 5) means that the values 1,2,3,4, and 5 have been tested.

*† recursive feature elimination includes iteratively removing the variable with the smallest Gini index.*

| **Score** | **Model** | **Feature selection** | **R^2^** | **MAE** |
| --- | --- | --- | --- | --- |
| *CAIDE* | *ElasticNet* | *None* | 0.45 | 1.57 |
|  | *Random Forest* | *Correlation-based* | 0.47 | 1.54 |
|  | *ElasticNet* | *Correlation-based* | 0.45 | 1.56 |
| *LIBRA* | *ElasticNet* | *None* | 0.01 | 1.30 |
|  | *Random Forest* | *Correlation-based* | 0.04 | 1.29 |
|  | *ElasticNet* | *Correlation-based* | 0.03 | 1.28 |

**SUPPLEMENTARY TABLE 10** **Performance of CAIDE and LIBRA prediction in the cross-validation of the EXTEND cohort.**

**SUPPLEMENTARY TABLE 11** **Performance of MCI or AD prediction in the independent test set of the EMIF-AD cohort.** Note that the value for which the geometric mean of the sensitivity and specificity is the highest in the cross-validation was used as a threshold for prediction.

| *Model* | *AUROC* | *TP* | *TN* | *FP* | *FN* | *MCC* | *Sensitivity* | *Specificity* | *PPV* | *NPV* | *Accuracy* | *Balanced accuracy* | *Cohen’s kappa* |
| --- | --- | --- | --- | --- | --- | --- | --- | --- | --- | --- | --- | --- | --- |
| MMRS-MCI (EN) | 0.6574 | 50 | 45 | 27 | 34 | 0.2196 | 0.5952 | 0.6250 | 0.6494 | 0.5696 | 0.6090 | 0.6101 | 0.2187 |
| MMRS-MCI (sPLS-DA) | 0.6486 | **53** | 45 | 27 | **31** | **0.2553** | **0.6310** | 0.6250 | **0.6625** | **0.5921** | **0.6282** | **0.6280** | **0.2549** |
| MMRS-MCI (RF-RFE) | **0.6847** | 44 | **49** | **23** | 40 | 0.2058 | 0.5238 | **0.6806** | 0.6567 | 0.5506 | 0.5962 | 0.6022 | 0.2010 |
| MMRS-AD  (EN) | 0.5444 | **15** | 45 | 27 | **16** | 0.1016 | **0.4839** | 0.6250 | 0.3571 | 0.7377 | 0.5825 | 0.5544 | 0.0989 |
| MMRS-AD (sPLS-DA) | 0.5470 | 14 | 48 | 24 | 17 | 0.1124 | 0.4516 | 0.6667 | 0.3684 | **0.7385** | 0.6019 | 0.5591 | 0.1111 |
| MMRS-AD (RF-RFE) | **0.6004** | 10 | **58** | **14** | 21 | **0.1390** | 0.3226 | **0.8056** | **0.4167** | 0.7342 | **0.6602** | **0.5641** | **0.1369** |
| Epi-CAIDE (MCI) | 0.5673 | 52 | **32** | **40** | 32 | 0.0643 | 0.6190 | **0.4444** | **0.5652** | 0.5000 | 0.5385 | **0.5317** | **0.064** |
| Epi-LIBRA (MCI) | **0.6134** | **61** | 24 | 48 | **23** | **0.0647** | **0.7262** | 0.3333 | 0.5596 | **0.5106** | **0.5449** | 0.5298 | 0.061 |
| Epi-CAIDE (AD) | 0.5233 | 16 | **37** | **35** | 15 | 0.0275 | 0.5161 | **0.5139** | **0.3137** | 0.7115 | **0.5146** | 0.5150 | **0.0254** |
| Epi-LIBRA (AD) | **0.5426** | **27** | 12 | 60 | **4** | **0.0477** | **0.8710** | 0.1667 | 0.3103 | **0.7500** | 0.3786 | **0.5188** | 0.0249 |
| PGSs  (EN) | 0.5967 | 48 | 40 | **32** | 36 | 0.1266 | 0.5714 | **0.5556** | 0.6000 | 0.5263 | 0.5641 | 0.5635 | 0.1265 |
| PGSs (sPLS-DA) | 0.6037 | 54 | **38** | 34 | 30 | 0.1715 | 0.6429 | 0.5278 | 0.6136 | 0.5588 | 0.5897 | 0.5853 | 0.1713 |
| PGSs (RF-RFE) | **0.6399** | **60** | **38** | 34 | **24** | **0.2466** | **0.7143** | 0.5278 | **0.6383** | **0.6129** | **0.6282** | **0.6210** | **0.2445** |
| MPSs/PGSs  (EN) | 0.6930 | 52 | 48 | 24 | 32 | **0.2850** | 0.6190 | 0.6667 | 0.6842 | **0.6000** | **0.6410** | **0.6429** | **0.2835** |
| MPSs/PGSs (sPLS-DA) | 0.6812 | 44 | **52** | **20** | 40 | 0.2494 | 0.5238 | **0.7222** | **0.6875** | 0.5652 | 0.6154 | 0.6230 | 0.2412 |
| MPSs/PGSs (RF-RFE) | **0.6935** | **53** | 46 | 26 | **31** | 0.2691 | **0.6310** | 0.6389 | 0.6709 | 0.5974 | 0.6346 | 0.6349 | 0.2685 |
| CSF  (EN) | 0.7634 | 55 | 17 | 5 | 28 | 0.3580 | 0.6627 | 0.7724 | 0.9167 | 0.3778 | 0.6857 | 0.7177 | 0.3145 |
| CSF  (sPLS-DA) | 0.7733 | 48 | **18** | **4** | 35 | 0.3227 | 0.5783 | **0.8182** | **0.9231** | 0.3396 | 0.6286 | 0.6982 | 0.2612 |
| CSF (RF-RFE) | **0.8097** | **60** | 16 | 6 | **23** | **0.3792** | **0.7229** | 0.7273 | 0.9091 | **0.4103** | **0.7238** | **0.7251** | **0.3506** |
| MPSs/CSF  (EN) | **0.8850** | 65 | 16 | 6 | 18 | 0.4439 | 0.7831 | 0.7273 | 0.9155 | 0.4706 | 0.7714 | 0.7552 | 0.4252 |
| MPSs/CSF (sPLS-DA) | 0.8631 | **69** | **17** | **5** | **14** | **0.5389** | **0.8313** | **0.7727** | **0.9324** | **0.5484** | **0.8190** | **0.8020** | **0.5251** |
| MPSs/CSF (RF-RFE) | 0.8614 | 62 | **17** | **5** | 21 | 0.4401 | 0.7470 | 0.7727 | 0.9254 | 0.4474 | 0.7524 | 0.7599 | 0.4101 |
| PGSs/CSF  (EN) | **0.7612** | 21 | 9 | 13 | 62 | -0.2939 | 0.2530 | 0.4091 | 0.6176 | 0.1268 | 0.2857 | 0.3311 | -0.1858 |
| PGSs/CSF (sPLS-DA) | 0.5865 | 0 | 22 | **0** | 83 | 0 | 0 | 1 | NaN | 0.2095 | 0.2095 | 0.5000 | 0 |
| PGSs/CSF (RF-RFE) | 0.7464 | **45** | 18 | 4 | **38** | **0.2940** | **0.5422** | **0.8182** | **0.9184** | **0.3214** | **0.6000** | **0.6802** | **0.2298** |
| MPSs/PGSs/CSF  (EN) | **0.8653** | 63 | 17 | 5 | 20 | 0.4530 | 0.7590 | 0.7727 | 0.9265 | 0.4595 | 0.7619 | 0.7659 | 0.4252 |
| MPSs/PGSs/CSF  (sPLS-DA) | 0.8631 | **69** | 14 | 8 | **14** | 0.4304 | **0.8313** | 0.6364 | 0.8961 | **0.5000** | **0.7905** | 0.7338 | 0.4251 |
| MPSs/PGSs/CSF  (RF-RFE) | 0.8401 | 62 | **18** | **4** | 21 | **0.4760** | 0.7470 | **0.8182** | **0.9294** | 0.4615 | 0.7619 | **0.7826** | **0.4402** |

Abbreviations: AUROC: area under the receiver operating characteristic curve; TP: true positives, TN: true negatives, FP: false positives, FN: false negatives, MCC: Matthews correlation coefficient, PPV: positive predicted value, NPV: negative predictive value.

**SUPPLEMENTARY TABLE 12** **The performance of risk factor prediction**. The performance was measured in the cross-validation of the EXTEND cohort for the different feature selection and machine learning methods.

| **Risk factor**  **(Performance measure)** | **Correlation, ElasticNet** | **Correlation, Random Forest** | **ElasticNet** | **Literature, ElasticNet** | **Literature, Random Forest** |
| --- | --- | --- | --- | --- | --- |
| *Education*  *(AUROC)* | **0.73** | 0.66 | 0.71 | 0.72 | 0.68 |
| *Physical inactivity*  *(AUROC)* | **0.61** | 0.58 | 0.57 | 0.54 | 0.52 |
| *Unhealthy diet*  *(AUROC)* | **0.63** | 0.57 | 0.54 | 0.54 | 0.56 |
| *Depression*  *(AUROC)* | **0.68** | 0.61 | 0.64 | 0.50 | 0.51 |
| *Type II Diabetes*  *(AUROC)* | **0.89** | 0.81 | 0.89 | 0.86 | 0.83 |
| *Heart Disease*  *(AUROC)* | **0.80** | 0.73 | 0.80 | 0.63 | 0.63 |
| *Sex*  *(AUROC)* | **1.00** | 1.00 | 1.00 | *NA* | *NA* |
| *Systolic blood pressure*  *(R^2^)* | 0.18 | 0.18 | **0.19** | 0.17 | 0.15 |
| *Total cholesterol*  *(R^2^)* | 2.6e-3 | **7.1e-3** | 2.6e-3 | 5.3e-3 | 3.5e-3 |

**SUPPLEMENTARY TABLE 13** **The performance of risk factor prediction in the EXTEND and EMIF-AD cohort.**

| **Variable type** | **Risk factor model** | **EXTEND** | **EMIF-AD** |
| --- | --- | --- | --- |
| *Continuous variables* | *Systolic blood pressure/hypertension* | R^2^ ≈ 0.19 * | AUROC ≈ 0.63 |
|  | *Total cholesterol* | R^2^ ≈ 7.1e-3 * | - |
|  | *Age* | R^2^ ≈ 0.92 | R^2^ ≈ 0.87 |
|  | *BMI/Obesity* | R^2^ ≈ 0.26 | AUROC ≈ 0.56 |
|  | *HDL cholesterol* | R^2^ ≈ 0.20 | - |
| *Discrete variables* | *Low education* | AUROC ≈ 0.73 * | AUROC ≈ 0.63 |
|  | *Physical inactivity* | AUROC ≈ 0.61 * | - |
|  | *Unhealthy diet* | AUROC ≈ 0.63 * | - |
|  | *Depression* | AUROC ≈ 0.68 * | AUROC ≈ 0.53 |
|  | *Type II diabetes* | AUROC ≈ 0.89 * | - |
|  | *Heart disease* | AUROC ≈ 0.80 * | AUROC ≈ 0.67 |
|  | *Sex* | AUROC = 1 * | AUROC = 1 |
|  | *Smoking* | AUROC ≈ 0.91 | AUROC ≈ 0.80 |
|  | *Alcohol consumption* | AUROC ≈ 0.57 | AUROC ≈ 0.58 |

^*^ Performance in the 5-repeated 5-fold cross-validation.

| **Cognitive outcome** | **Log-rank test p-value** | **Hazard ratio**  **(95% C.I)** | **Cox regression**  **p-value** | **AUROC**  **(95 % C.I.)** | **N *** |
| --- | --- | --- | --- | --- | --- |
| *MMSE* | 0.3 | 1.56 (0.64;3.82) | 0.3 | 0.57 (0.42;0.72) | 56/58/58 |
| *RAVLT- Learning* | **0.02** | 2.11 (1.08;4.11) | **0.028** | 0.62 (0.51;0.72) | 59/60/61 |
| *RAVLT – Forgetting* | 0.5 | 1.21 (0.65;2.25) | 0.6 | 0.53 (0.41;0.65) | 55/58/55 |
| *RAVLT – Percent forgetting* | 0.2 | 1.36 (0.87;2.15) | 0.2 | 0.57 (0.46;0.67) | 61/61/66 |
| *RAVLT – Immediate recall* | 0.3 | 1.39 (0.78;2.48) | 0.3 | 0.56 (0.45;0.66) | 59/60/61 |
| *ADAS-Q4* | 0.2 | 1.52 (0.85;2.72) | 0.2 | 0.59 (0.48;0.69) | 58/61/61 |
| *ADAS13* | 0.2 | 1.44 (0.83;2.50) | 0.2 | 0.57 (0.47;0.67) | 62/60/62 |
| *ADAS11* | 0.3 | 1.28 (0.77;2.12) | 0.3 | 0.58 (0.48;0.68) | 62/62/63 |
| *LDELTOTAL* | 0.4 | 1.26 (0.72;2.18) | 0.4 | 0.55 (0.44;0.65) | 61/60/64 |
| *TRABSCOR* | **0.04** | 1.94 (1.01;3.73) | **0.048** | 0.60 (0.49;0.71) | 60/63/62 |

**SUPPLEMENTARY TABLE 14 Statistics of survival analysis for the different cognitive outcomes in the ADNI cohort.**

* The number of samples in the low/intermediate/high risk group.

**SUPPLEMENTARY TABLE 15 Top 20 most significantly enriched GO (BP) terms.**

| **GO ID** | **Description** | **Total # Genes** | **# genes in MMRS-MCI model** | **p-value** | **FDR-adjusted p-value** |
| --- | --- | --- | --- | --- | --- |
| *GO:0097113* | *AMPA glutamate receptor clustering* | 9 | 8 | 0.000416 | 1 |
| *GO:0097688* | *glutamate receptor clustering* | 9 | 8 | 0.000416 | 1 |
| *GO:0044794* | *positive regulation by host of viral process* | 13 | 9 | 0.00062 | 1 |
| *GO:0072683* | *T cell extravasation* | 10 | 7 | 0.000659 | 1 |
| *GO:0097435* | *supramolecular fiber organization* | 705 | 227 | 0.000808 | 1 |
| *GO:0007417* | *central nervous system development* | 973 | 308 | 0.000915 | 1 |
| *GO:0007229* | *integrin-mediated signaling pathway* | 104 | 45 | 0.001104 | 1 |
| *GO:0048706* | *embryonic skeletal system development* | 119 | 49 | 0.001431 | 1 |
| *GO:0044788* | *modulation by host of viral process* | 26 | 13 | 0.001447 | 1 |
| *GO:0048705* | *skeletal system morphogenesis* | 217 | 81 | 0.001449 | 1 |
| *GO:0010810* | *regulation of cell-substrate adhesion* | 213 | 80 | 0.001499 | 1 |
| *GO:0007010* | *cytoskeleton organization* | 1359 | 405 | 0.002014 | 1 |
| *GO:0022604* | *regulation of cell morphogenesis* | 295 | 107 | 0.00221 | 1 |
| *GO:0045785* | *positive regulation of cell adhesion* | 423 | 134 | 0.002367 | 1 |
| *GO:0007188* | *adenylate cyclase-modulating G protein-coupled receptor signaling pathway* | 226 | 73 | 0.002642 | 1 |
| *GO:0002861* | *regulation of inflammatory response to antigenic stimulus* | 41 | 18 | 0.002738 | 1 |
| *GO:1903724* | *positive regulation of centriole elongation* | 4 | 4 | 0.00302 | 1 |
| *GO:0050804* | *modulation of chemical synaptic transmission* | 424 | 146 | 0.003386 | 1 |
| *GO:0072578* | *neurotransmitter-gated ion channel clustering* | 13 | 9 | 0.003576 | 1 |
| *GO:0001913* | *T cell mediated cytotoxicity* | 46 | 17 | 0.003587 | 1 |

**SUPPLEMENTARY TABLE 16 The performance of risk factor prediction by the PGSs in the EXTEND and EMIF-AD cohort.**

| **Variable type** | **Risk factor model** | **EXTEND** | **EMIF-AD** |
| --- | --- | --- | --- |
| *Continuous variables* | *Systolic blood pressure/hypertension* | R^2^ ≈ 0.073 | AUROC ≈ 0.65 |
|  | *Total cholesterol* | R^2^ ≈ 0.072 | - |
|  | *BMI/Obesity* | R^2^ ≈ 0.091 | AUROC ≈ 0.67 |
|  | *HDL cholesterol* | R^2^ ≈ 0.10 | - |
| *Discrete variables* | *Low education* | AUROC ≈ 0.66 | AUROC ≈ 0.60 |
|  | *Physical inactivity* | AUROC ≈ 0.52 | - |
|  | *Unhealthy diet* | AUROC ≈ 0.56 | - |
|  | *Depression* | AUROC ≈ 0.54 | AUROC ≈ 0.45 |
|  | *Type II diabetes* | AUROC ≈ 0.60 | - |
|  | *Heart disease* | AUROC ≈ 0.62 | AUROC ≈ 0.48 |
|  | *Alcohol consumption* | AUROC ≈ 0.54 | AUROC ≈ 0.48 |

### Supplementary Figures

| **(A)** | ***ADAS11*** |
| --- | --- |
|  | 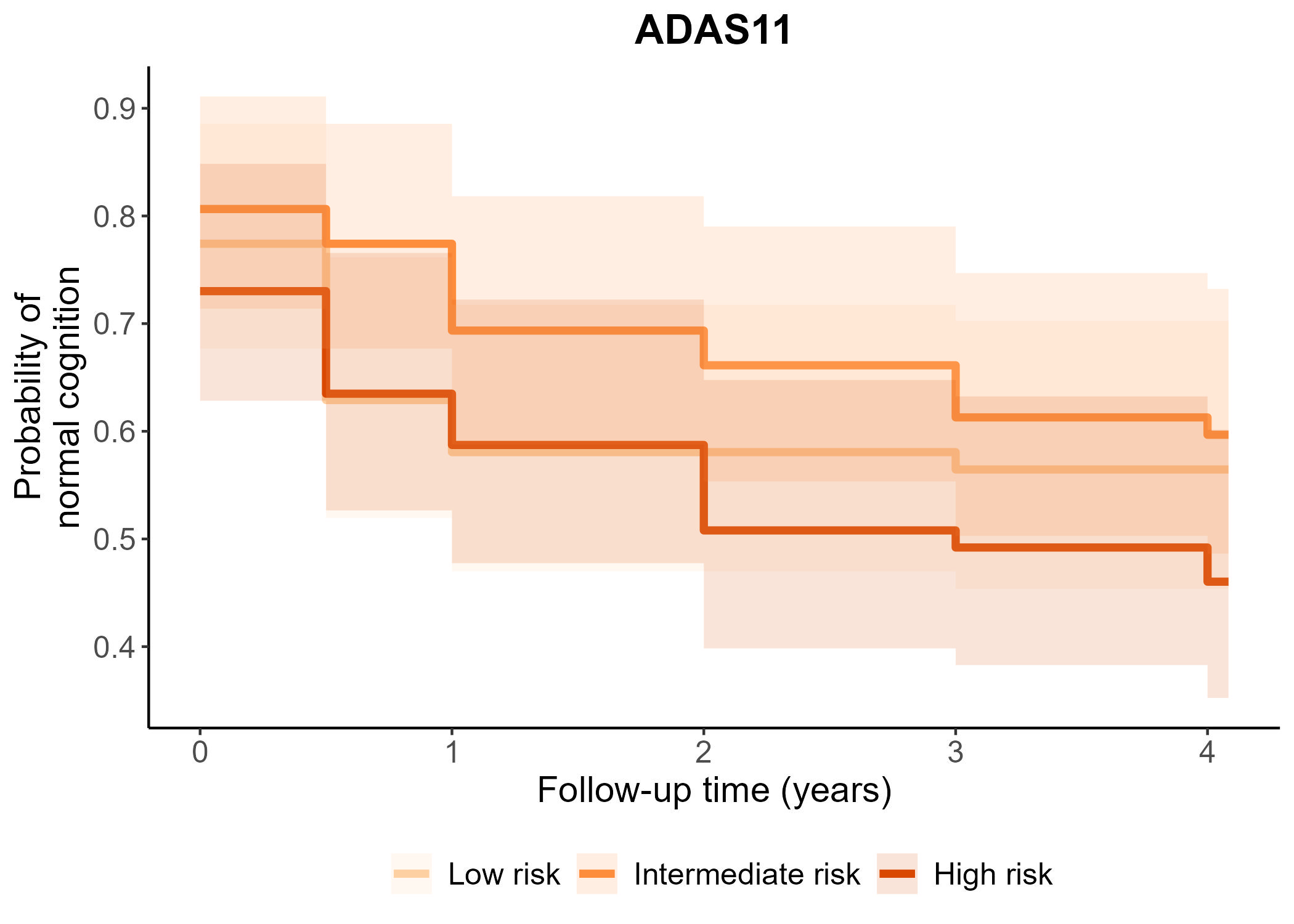 |
| **(B)** | **ADAS13** |
|  | 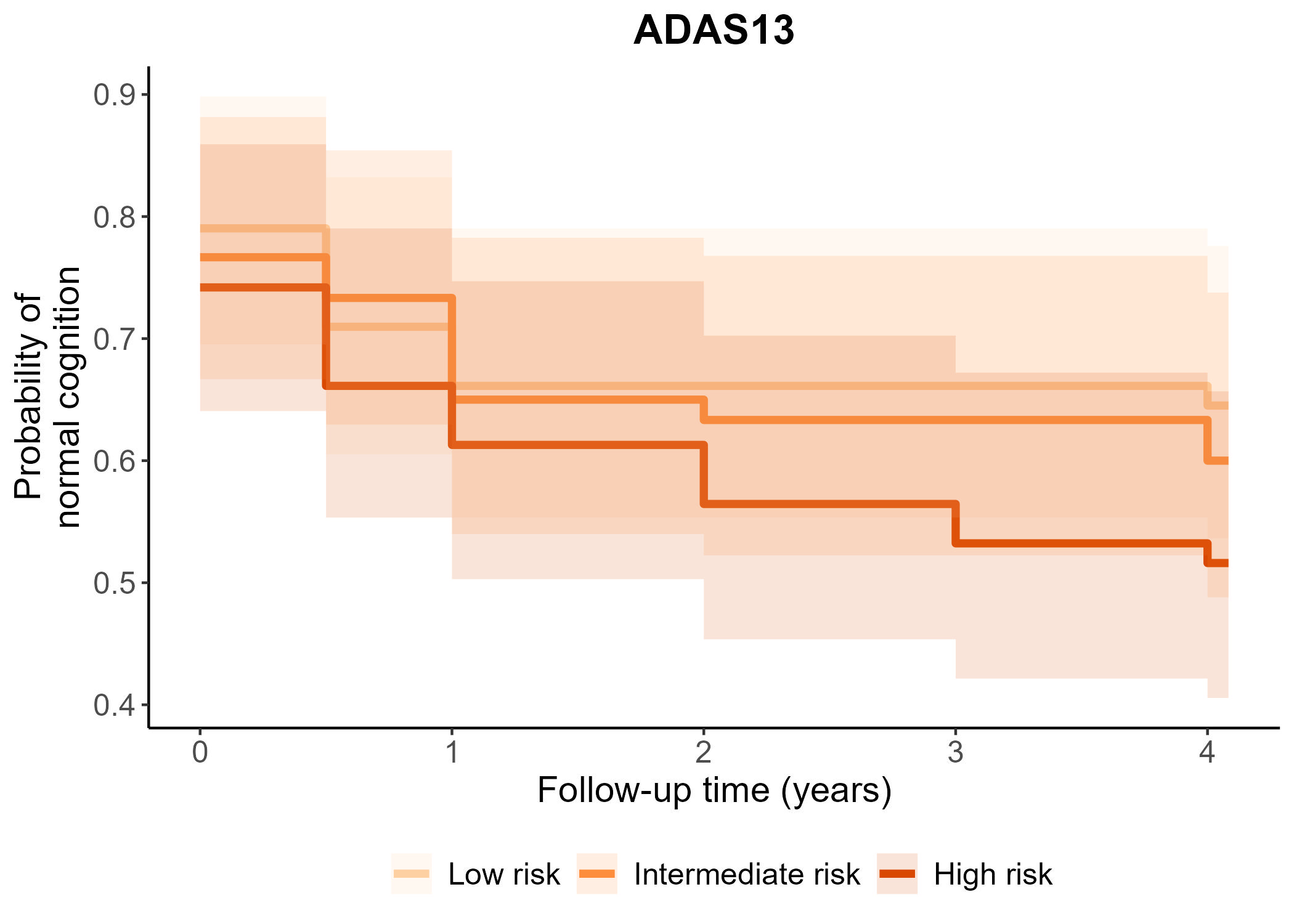 |
| **(C)** | **ADAS-Q4** |
|  | 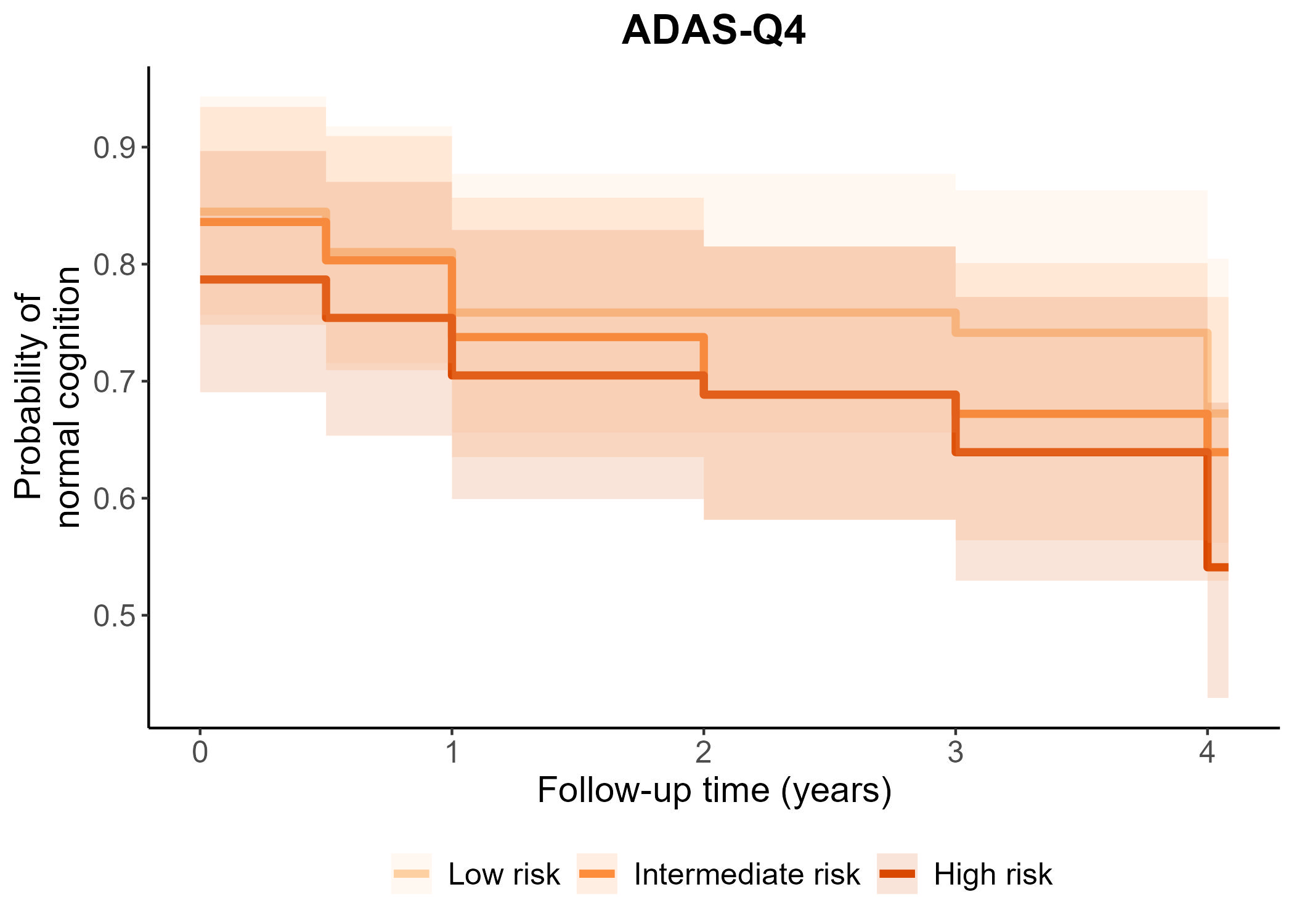 |
| **(D)** | **LDELTOTAL** |
|  | 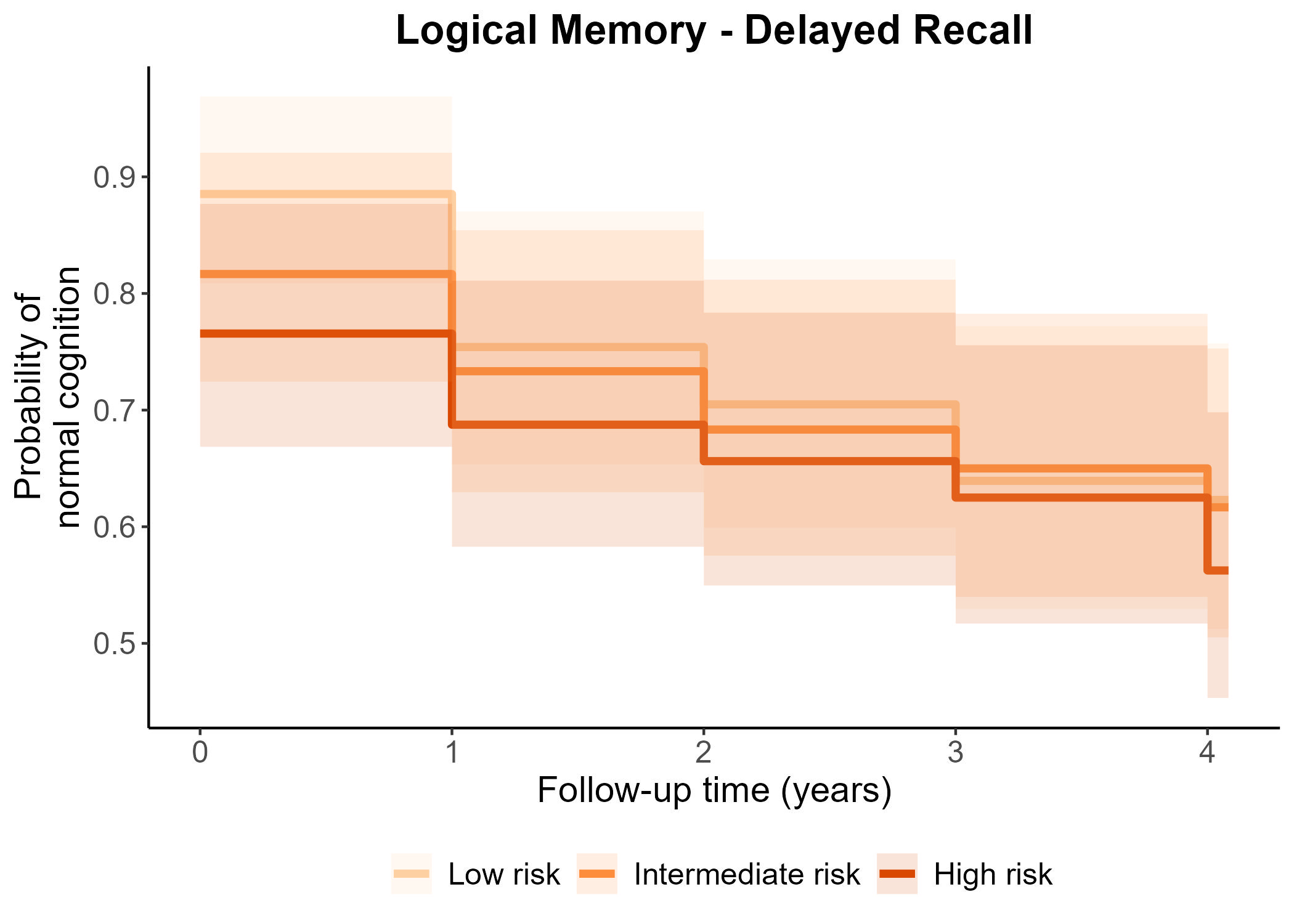 |
| **(E)** | **MMSE** |
|  | 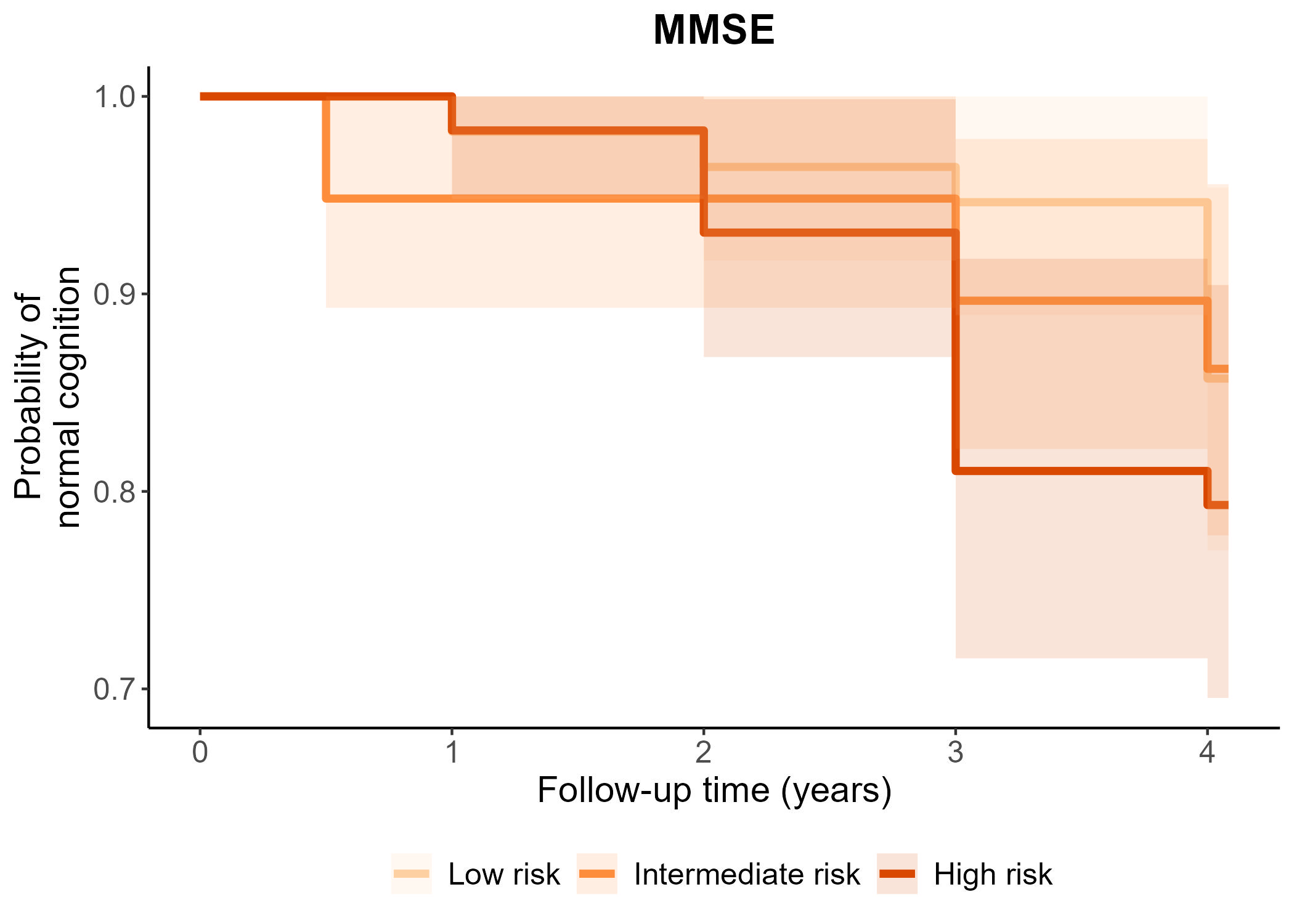 |
| **(F)** | **RAVLT – Forgetting** |
|  | 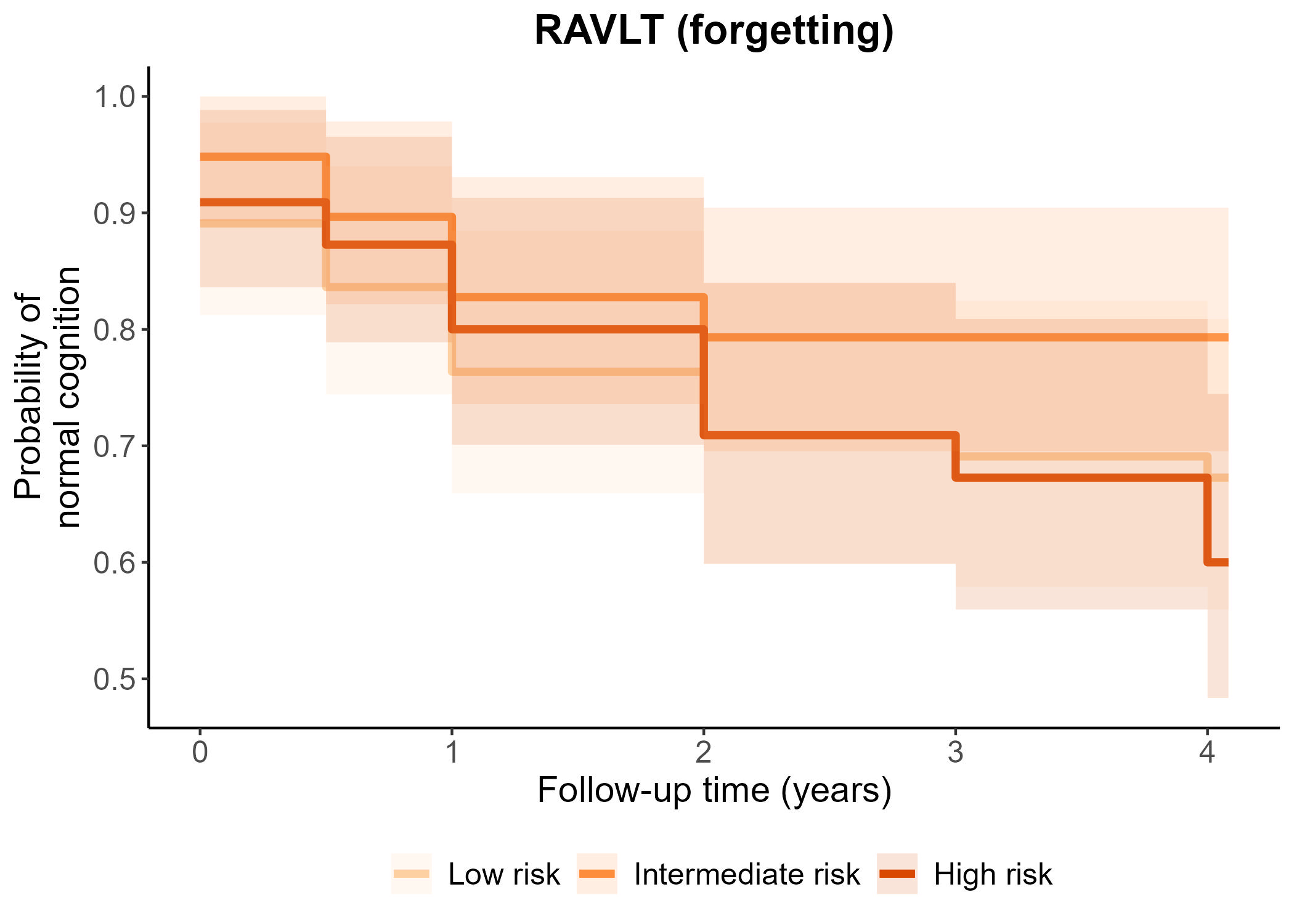 |
| **(G)** | **RAVLT – Percent forgetting** |
|  | 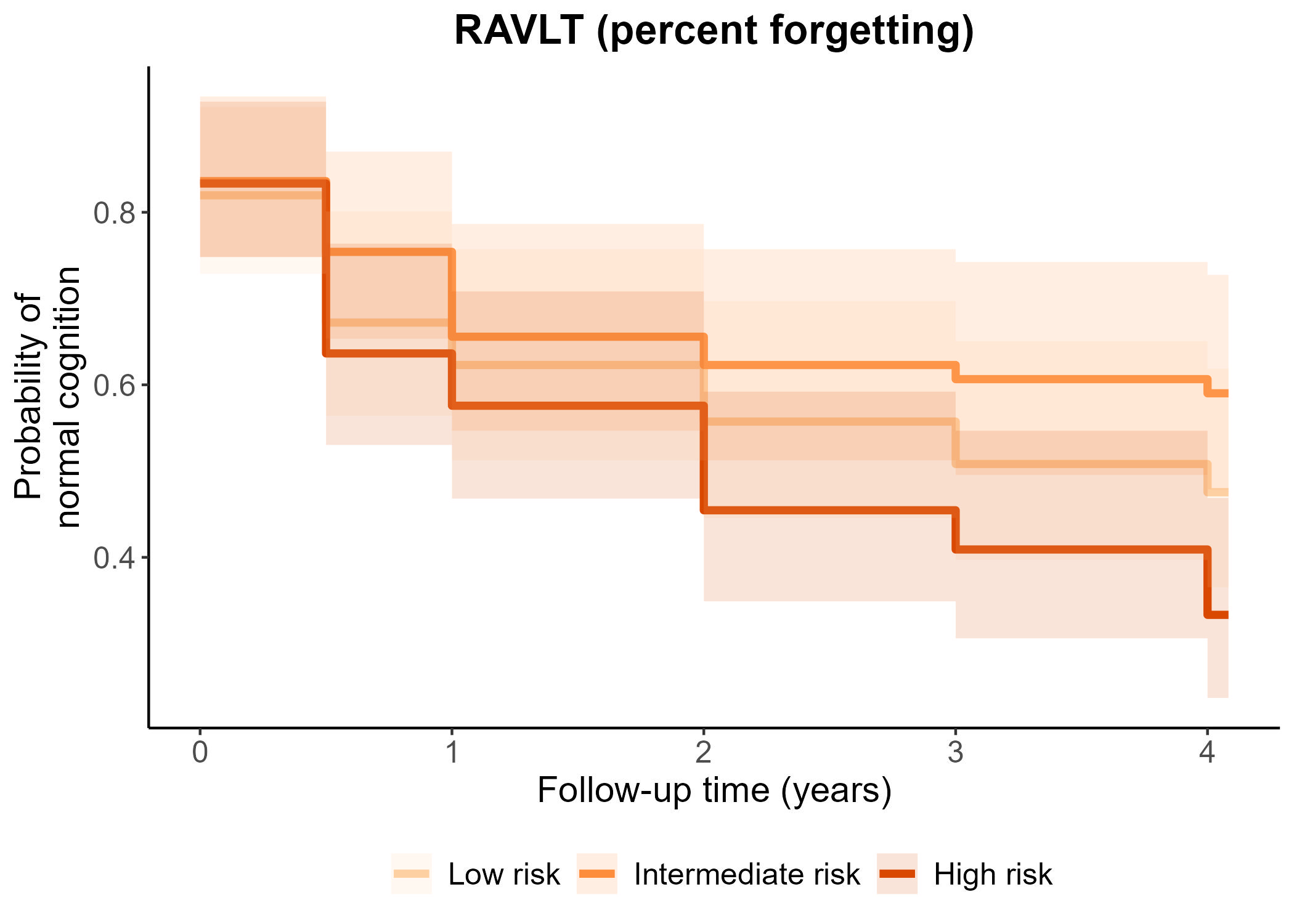 |
| **(H)** | **RAVLT – Immediate recall** |
|  | 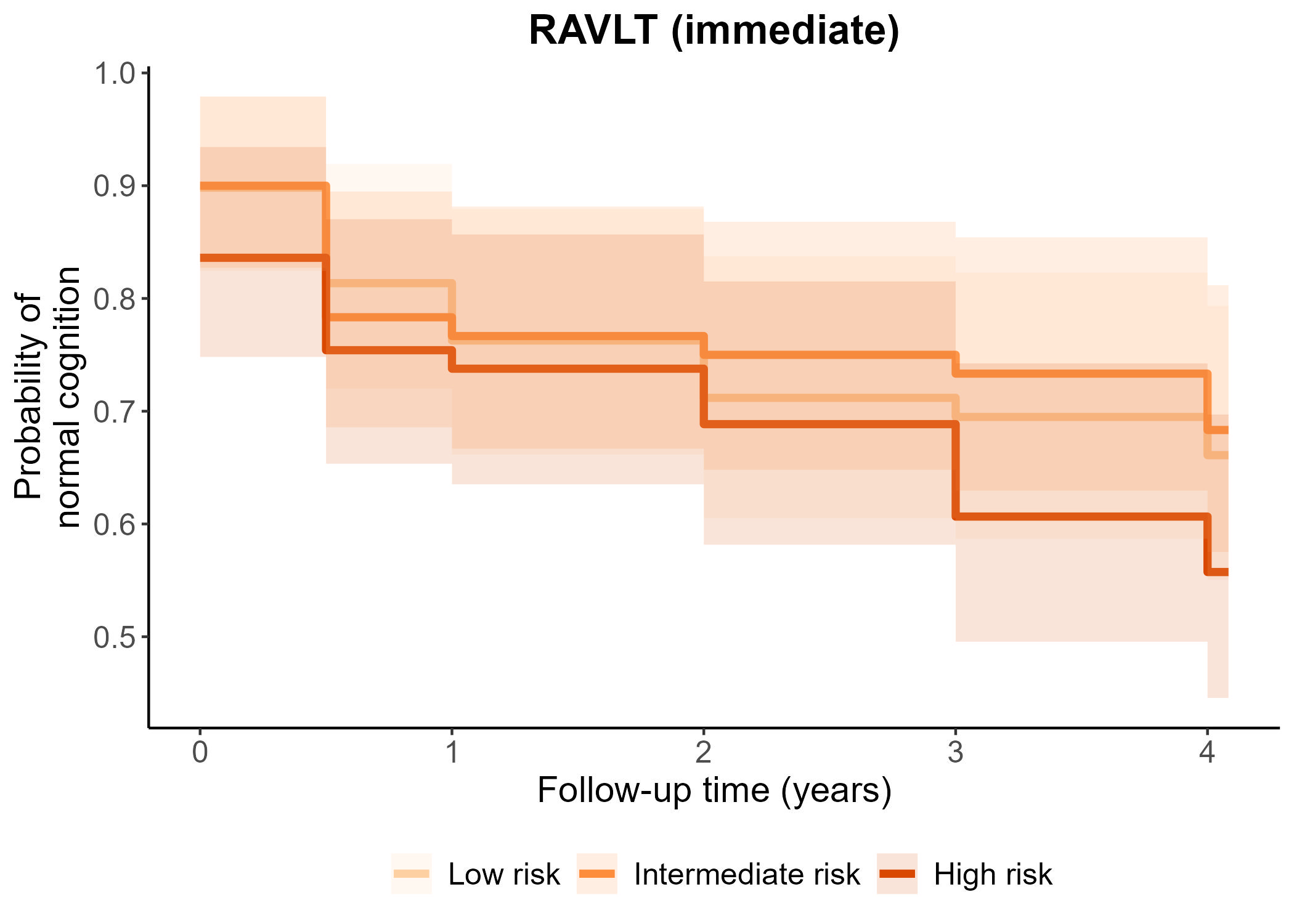 |
| **(I)** | **RAVLT - Learning** |
|  | 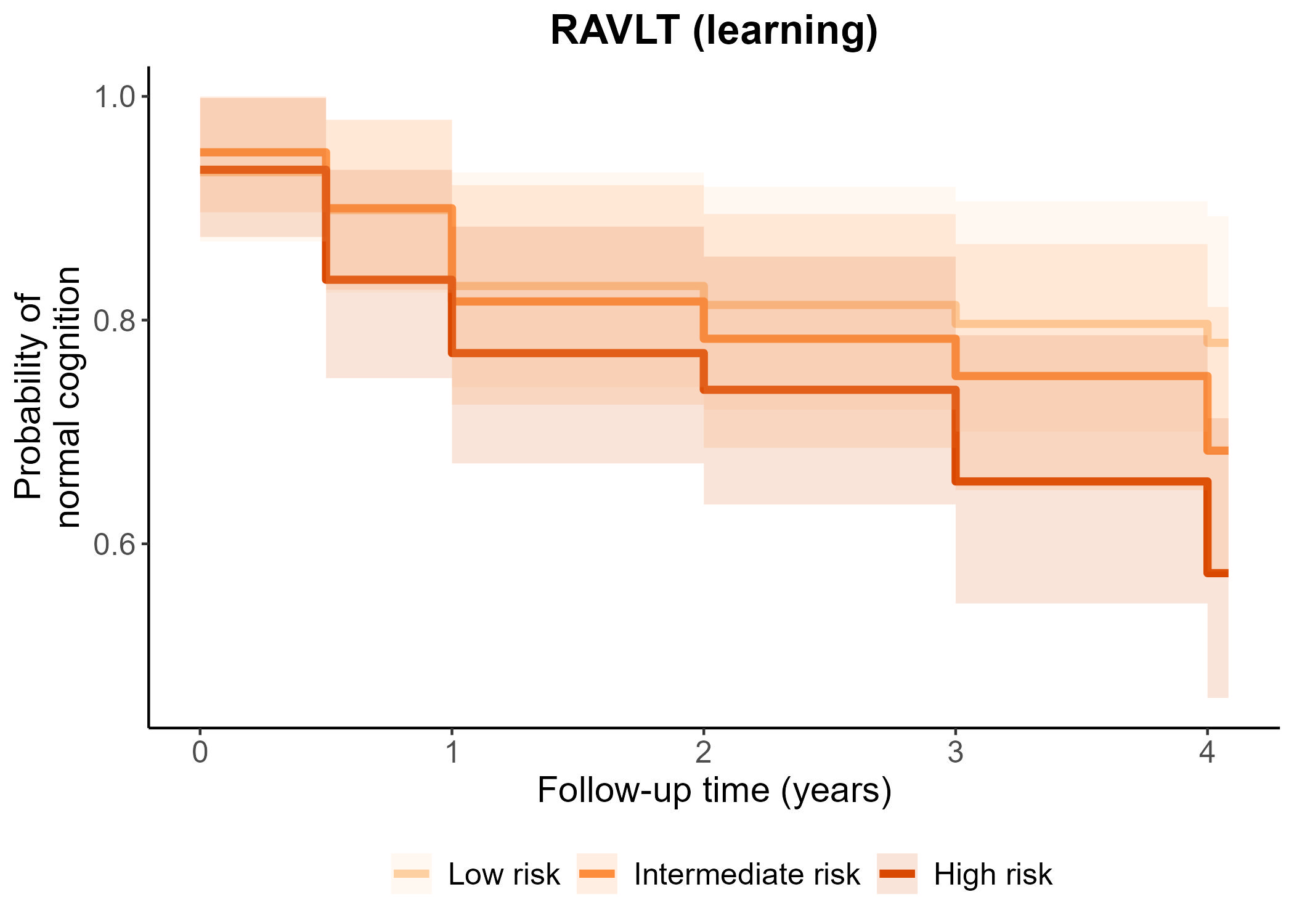 |
| **(J)** | **TRABSCOR** |
|  | 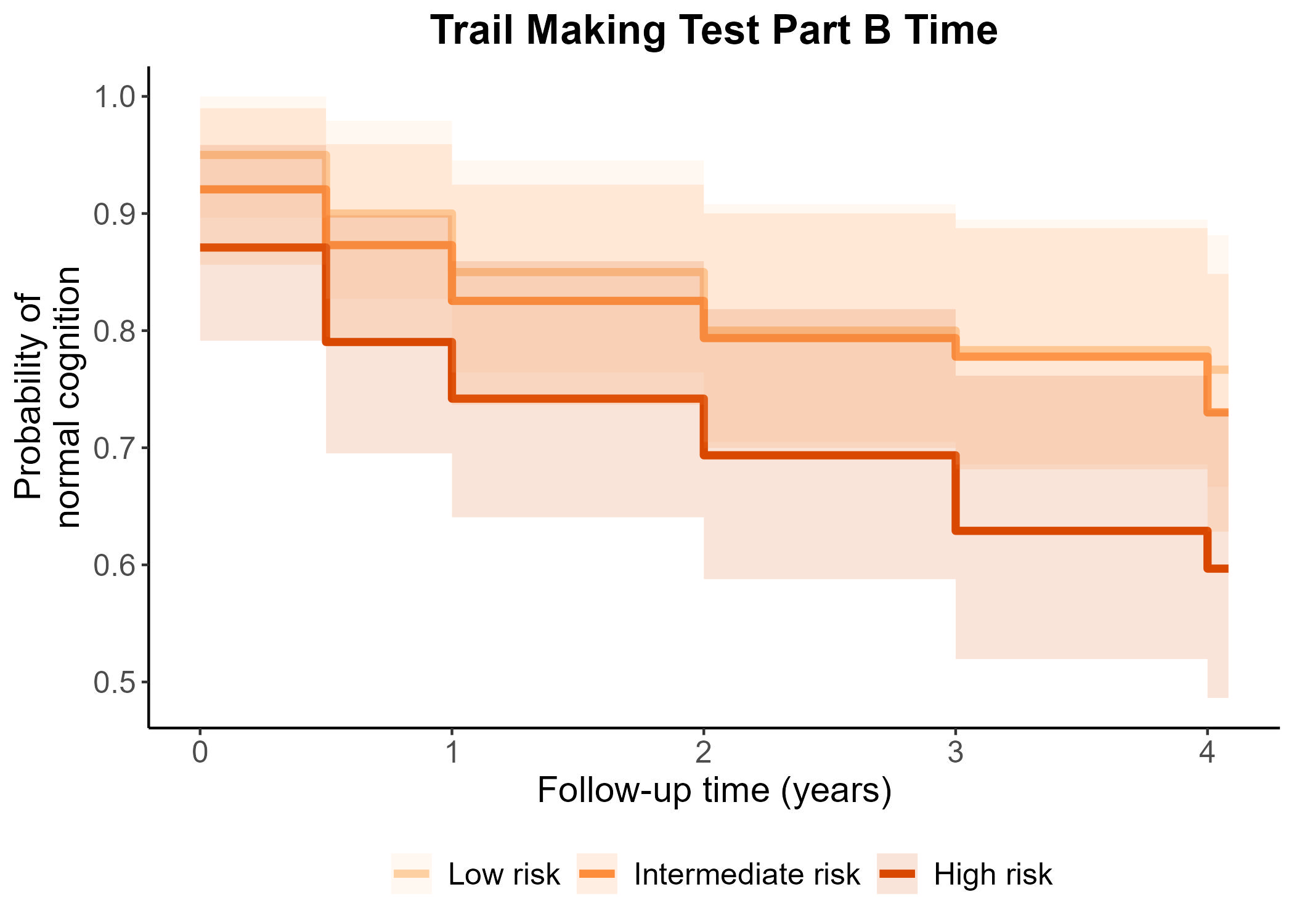 |

**SUPPLEMENTARY FIGURE 1** Kaplan-Meier curves of cognitive impairments according to difference cognitive tests in the ADNI cohorts.

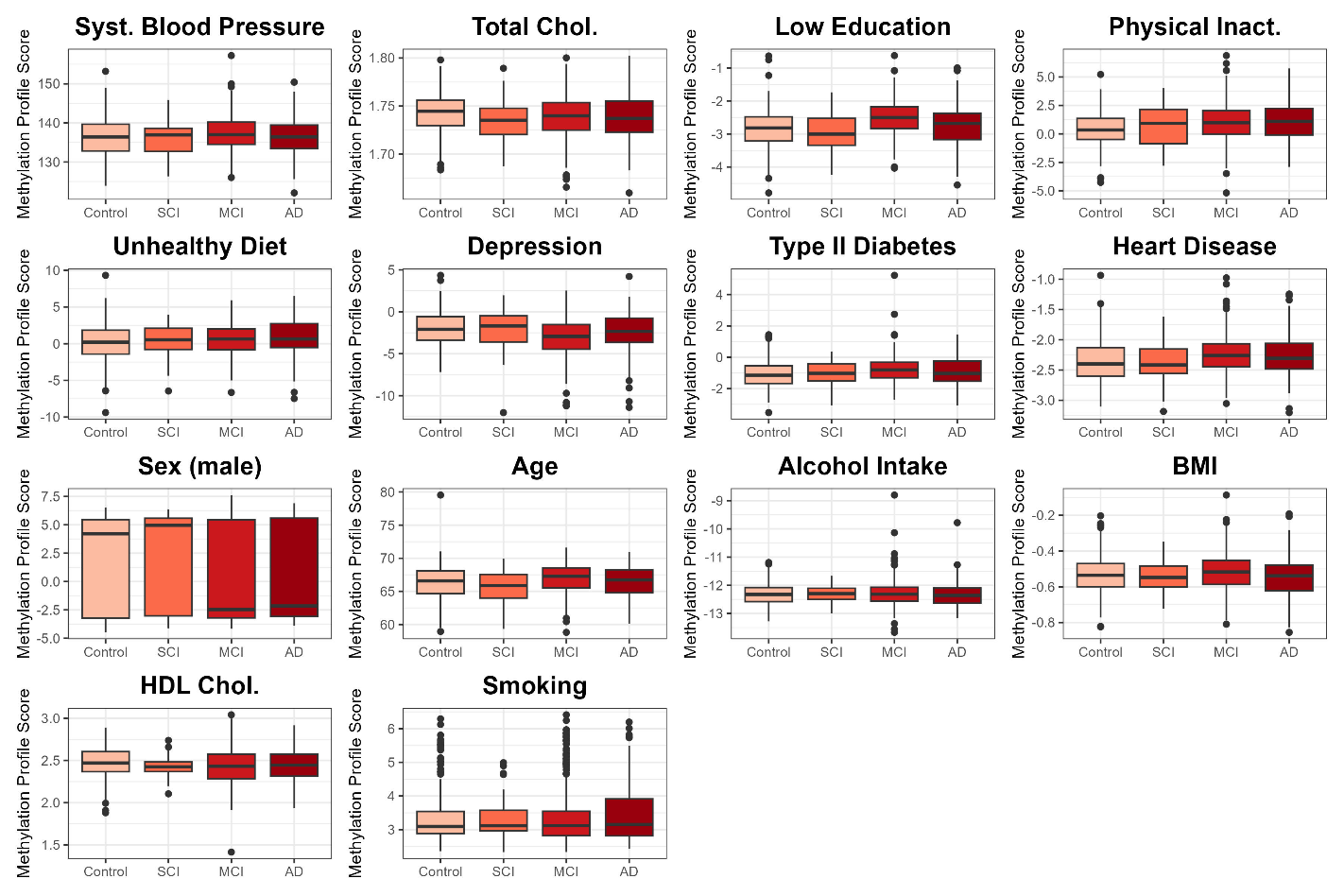

**SUPPLEMENTARY FIGURE 2** Boxplots of the 14 MPSs for the four different diagnostic groups. Abbreviations: SCI: subjective cognitive impairments, MCI: mild cognitive impairments, AD: Alzheimer’s disease.

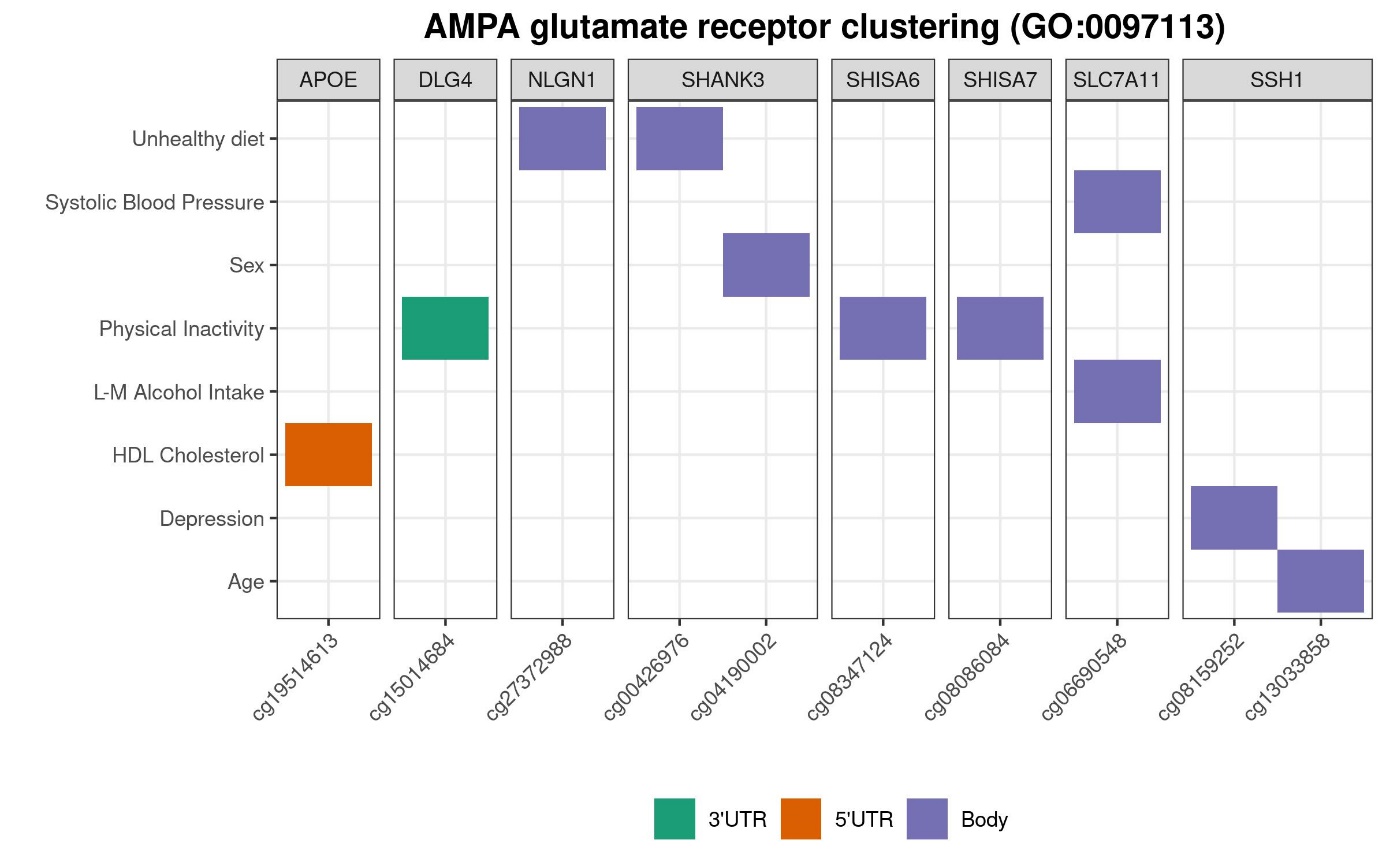

**SUPPLEMENTARY FIGURE 3** CpGs of the dementia risk factor models associated with *the AMPA glutamate receptor clustering* GO term.

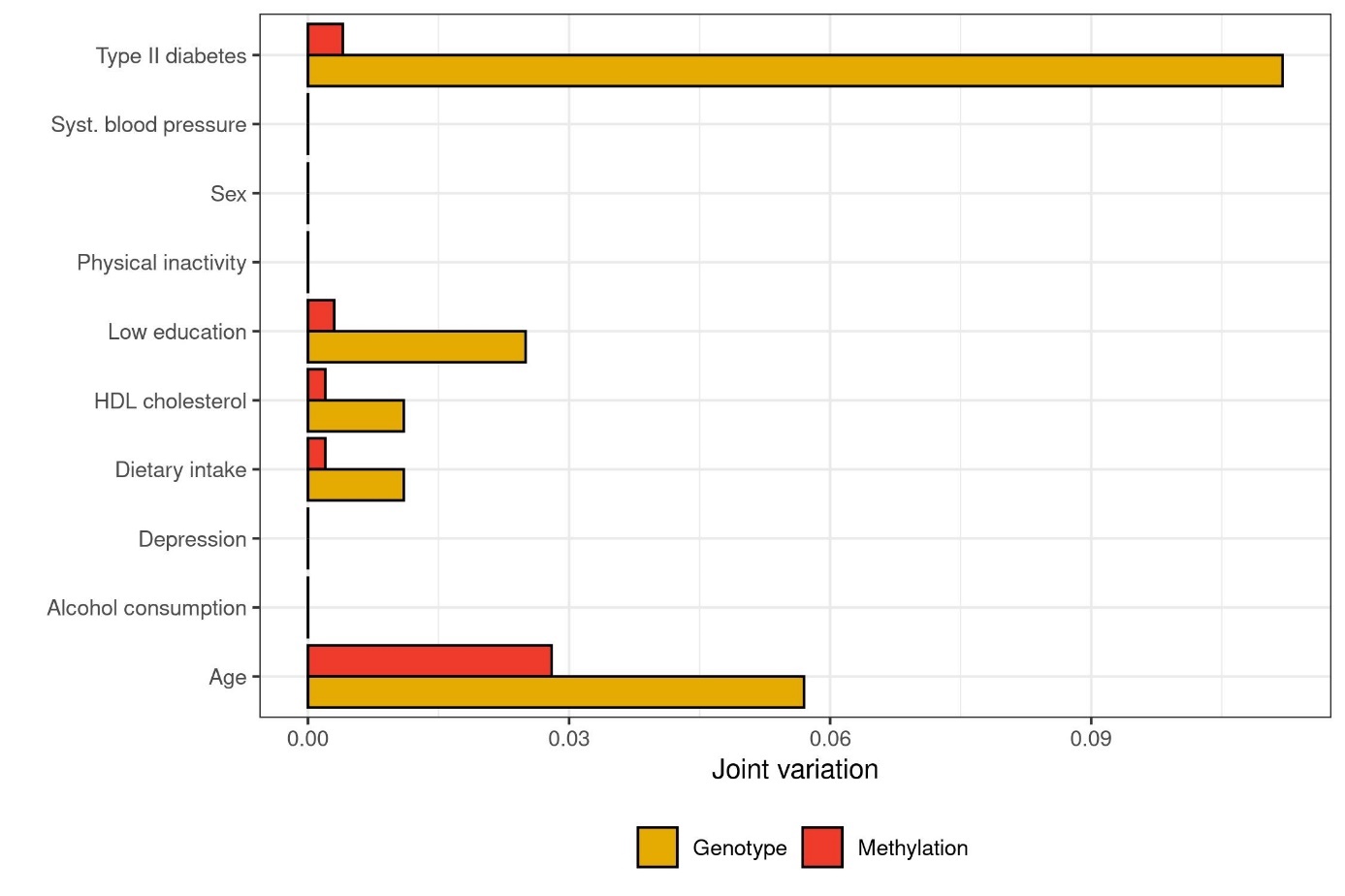
**SUPPLEMENTARY FIGURE 4** Joint variation (JIVE) of the model’s CpGs (methylation) and their mQTLs (genotype). The analysis was only performed on the risk factor model that are included in the MMRS-MCI (RF-RFE) model.

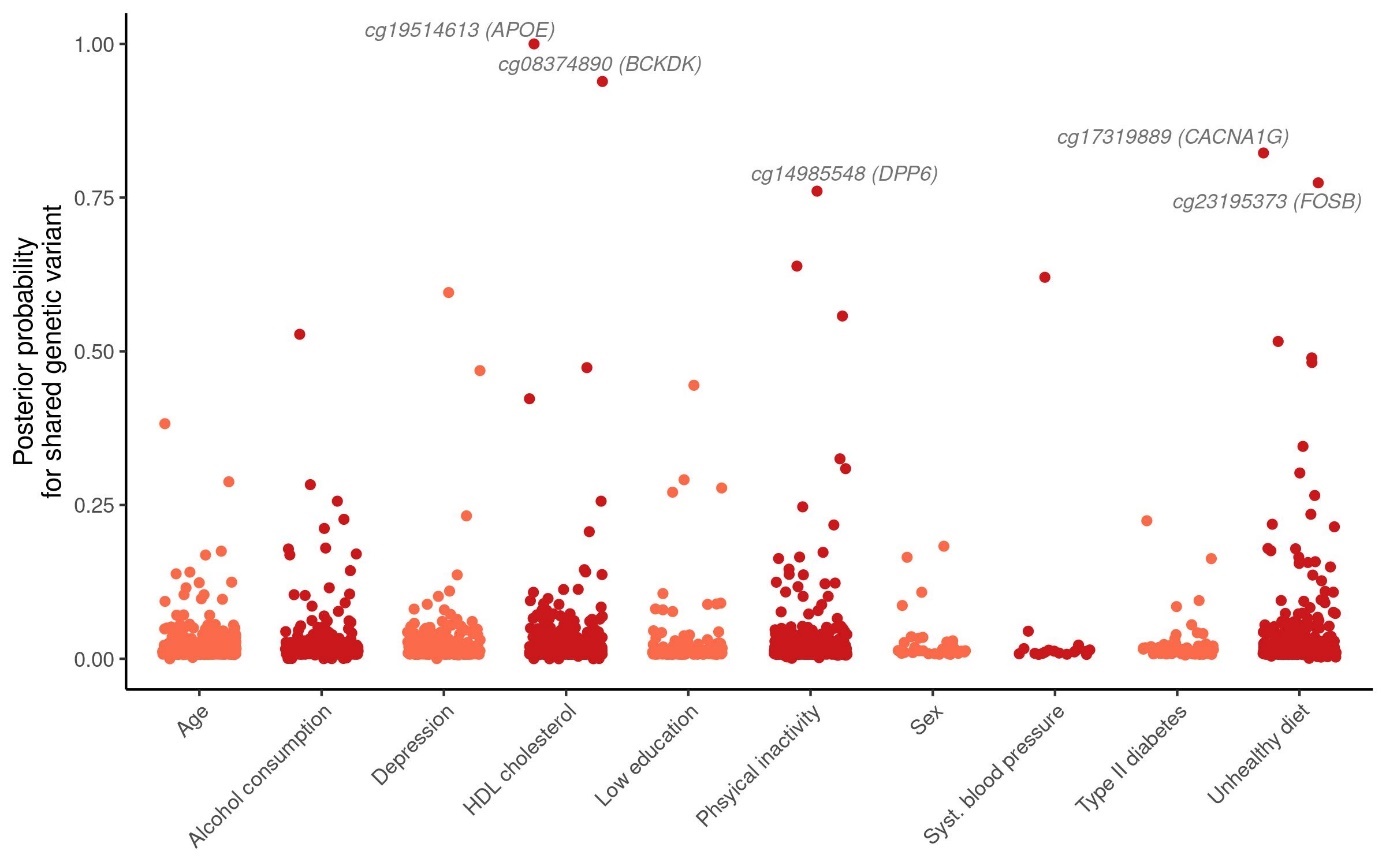

**SUPPLEMENTARY FIGURE 5** Posterior probability of the model’s CpGs for having a shared genetic variant with the AD status (*i.e.,* hypothesis 4 of the Bayes Factor colocalization analysis).

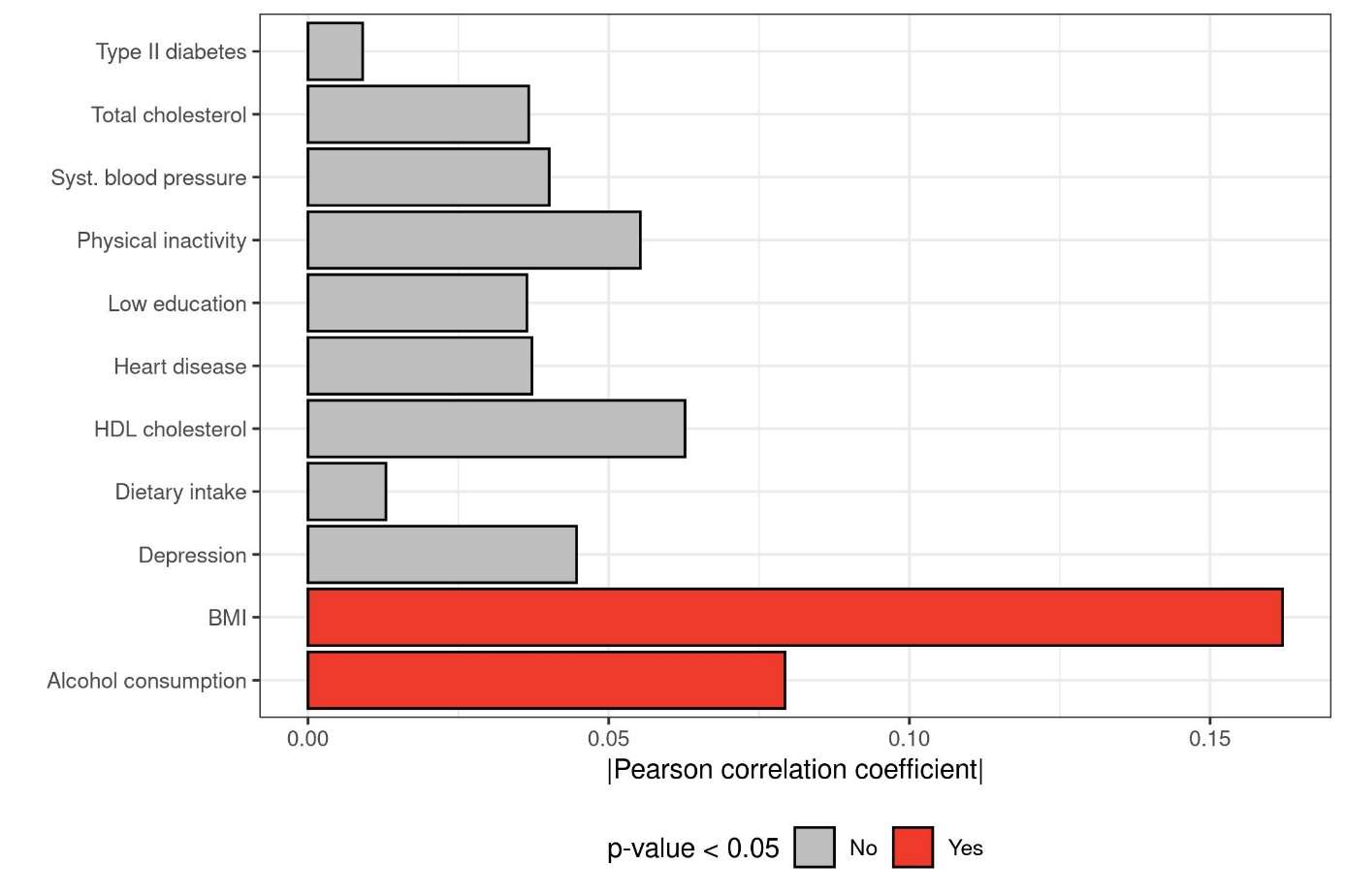

**SUPPLEMENTARY FIGURE 6** Correlations between corresponding PGSs and MPSs.

### Supplementary Texts

**Supplementary Text 1**

**Supplementary Methods**

To make the machine learning approaches computationally tractable, feature selection can be performed to remove less influential variables and reduce the dimensionality of the data. Therefore, various supervised and unsupervised feature selection methods were evaluated in terms of the predictive performance of an ElasticNet-regularized linear regression model in predicting the CAIDE score. For this, the same data preprocessing of the DNA methylation data was performed as described in the main text. However, before the evaluation of the feature selection methods, the EXTEND data was divided into a training (n = 924) and independent test (n = 152) set using the Kennard-Stone algorithm [22].

*Variance-based Feature Selection*

Variance-based feature selection approaches are commonly used to deal with the large dimensionality of DNA methylation data [23]. In this approach, the top features with the highest variance are selected. In the present study, four different approaches to this feature selection method were evaluated. Specifically, in the first two approaches, the 10,000 probes with the most variable β- and M-values were selected for model training. These methods will be referred to as variance (β) and variance (M) feature selection methods. Furthermore, in the third and fourth approaches, referred to as variance (β, Cor) and variance (M, Cor) approaches, cell type composition (as estimated by the *wateRmelon* package [24]) was regressed out before selecting the 10,000 features with the most variable β- and M-values. Noteworthily, in this approach, the adjusted β- and M-values were only used to determine which probes are the most variable, while the unadjusted β- and M-values of the selected probes were used in the subsequent analysis (e.g., prediction of CAIDE1 score).

*S-score-based Feature Selection*

The next feature selection method that was evaluated included the selection of the 10,000 features with the highest S-score (**Supplementary Equation 1**). This S-score-based approach preferentially selects low-variance features with a mean β-value close to zero or one and has been developed by Gerritse [25] to deal with phenotypes with less pronounced alterations.

$$S= \frac{|mean\left( \beta\right)- 0.5|}{Var(\beta)} \mathbf{(Supplementary Equation 1)}$$

*PCA-based Feature Selection*

For the PCA-based feature selection, PCA was performed on the whole training data (n = 924). All 924 principal components were accordingly used as features for the model training and validation. The PCA scores of the test set were calculated through matrix multiplication of the β-values with the loadings of the established PCA model.

*Kennard-Stone-like Feature Selection*

The Kennard-Stone algorithm is a commonly used method for the selection of a representative sample subset from the data. This algorithm works by first selecting the two most distant samples and afterwards iteratively adding the most distant sample to the sample set [22]. Interestingly, by iteratively adding the most uncorrelated probe to the feature set, the Kennard-Stone algorithm could potentially be used to select a representative feature set from the data.

To reduce computational time, an adjusted version of the Kennard-Stone algorithm was implemented for the feature selection (**Supplementary Fig. 7**). Specifically, instead of starting with the two most distant probes, in the current implementation, the Kennard-Stone selection started with the probe with the highest variance (*i.e.,* seed probe) after which the probes with the smallest maximal absolute Spearman correlation to all selected probes was added to the feature set. As a consequence of starting with a seed probe, not all possible pairwise comparisons needed to be calculated, thus decreasing the computational time significantly.

In addition, the features were divided into 54 subsections based on biological and technical probe characteristics. These characteristics include the probe type (*i.e.,* type I and type II), the location of the CpG with respect to the genes (*i.e.,* intergenic, promotor, gene body) and the CpG islands (*i.e.,* shore/shelf, island, open sea), and the variance of the probe (i.e., low, intermediate, high variance). Performing the feature selection separately for the different subsections reduces the number of pairwise comparisons and allows for the implementation of parallel computing, thereby significantly decreasing the computational complexity. To ensure that the final features are a representative set with limited overlapping information, selected features from each subsection were combined and a final Kennard-Stone feature selection round was used to get to the final feature set. Due to these adjustments, the feature selection method will be referred to as Kennard-Stone-like (KS-like) feature selection.

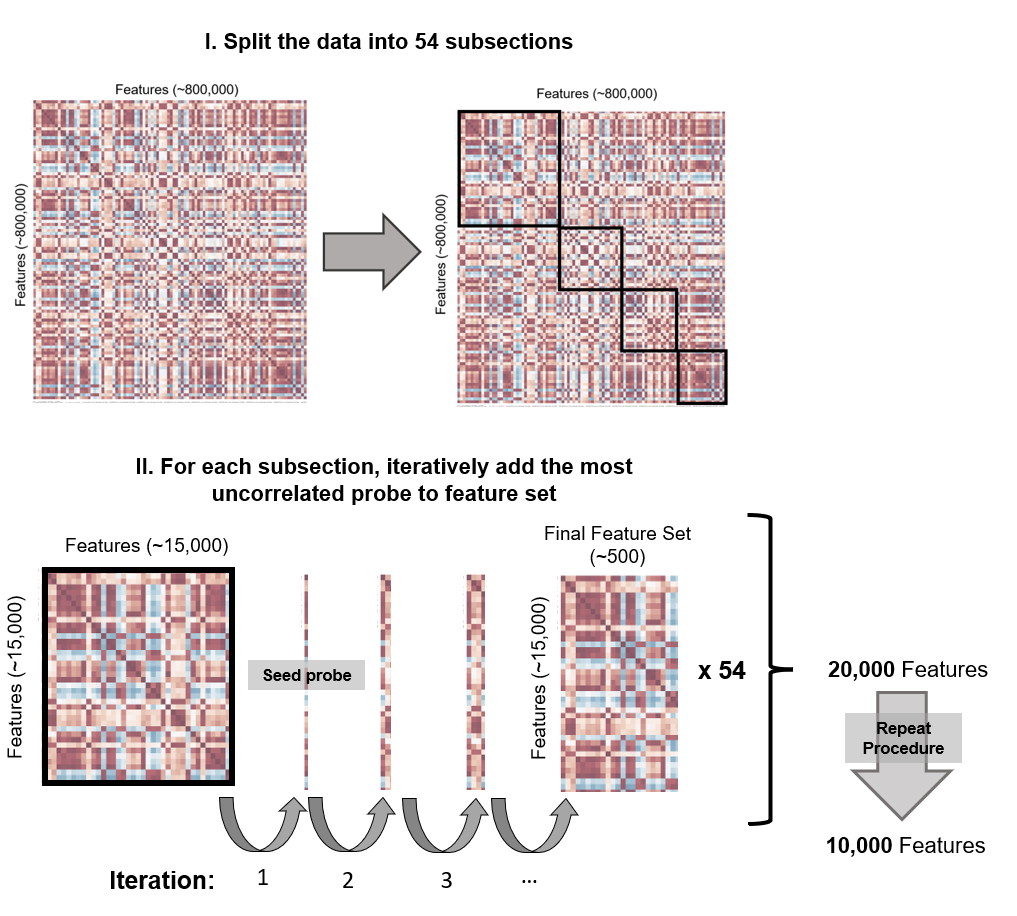

**SUPPLEMENTARY FIGURE 7** Overview of Kennard-Stone (KS)-like feature selection. In the first step of the feature selection, the data is split into 54 subsections based on the probe type, location of CpG, and feature variance. Subsequently, for each subsection, starting from the probe with the highest variance (*i.e.,* seed probe), the most uncorrelated probe is iteratively added to the feature set. A final feature selection round is performed to get to the final 10,000 features. Note that the heatmap in the figure is only meant for visualization purposes, the dimensions and colours are not representative of the real DNA methylation data.

*Correlation-based Feature Selection*

In the correlation-based feature selection method, the Spearman correlations between the probe’s β-values and the target variable (*i.e.,* LIBRA, CAIDE, or dementia risk factors) were calculated and the 10,000 probes with the highest absolute Spearman correlation were selected as variables for the prediction.

However, for the prediction of the CAIDE and LIBRA scores, the same number of top correlated probes were selected for each of the included risk factors (**Supplementary Table 2 and 3**) until a combined number of 10,000 selected variables. By selecting the same number of top correlated probes for each of the included risk factors, the current approach avoids that (almost) all selected probes relate to only a single risk factor such as age or sex. Noteworthily, since this is a supervised approach, the feature selection is performed within the cross-validation loop.

To enhance computational efficiency, the feature selection within the cross-validation was not performed on all (approx. 800,000) features, but instead on the 70,000 probes that have the highest absolute Spearman correlation with the target variable (*i.e.,* predicted risk factor) in the whole EXTEND data. For the prediction of the LIBRA and CAIDE scores, however, the feature selection was performed on the union of the 10,000 most correlated features of the individual risk factors in the whole training data (*i.e.,* 57,881 probes for CAIDE and 98,099 probes for LIBRA).

**Supplementary Results**

**Supplementary Fig. 8** shows the root mean squared error (RMSE) of the optimized ElasticNet-regularized linear regression model in the cross-validation and test set for each feature selection approach. In this figure, it can be seen that the ElasticNet model with prior correlation-based feature selection outperformed all unsupervised feature selection methods. In addition, despite being computationally more efficient, the correlation-based approach predicted the CAIDE score even slightly better than the model without prior feature selection.

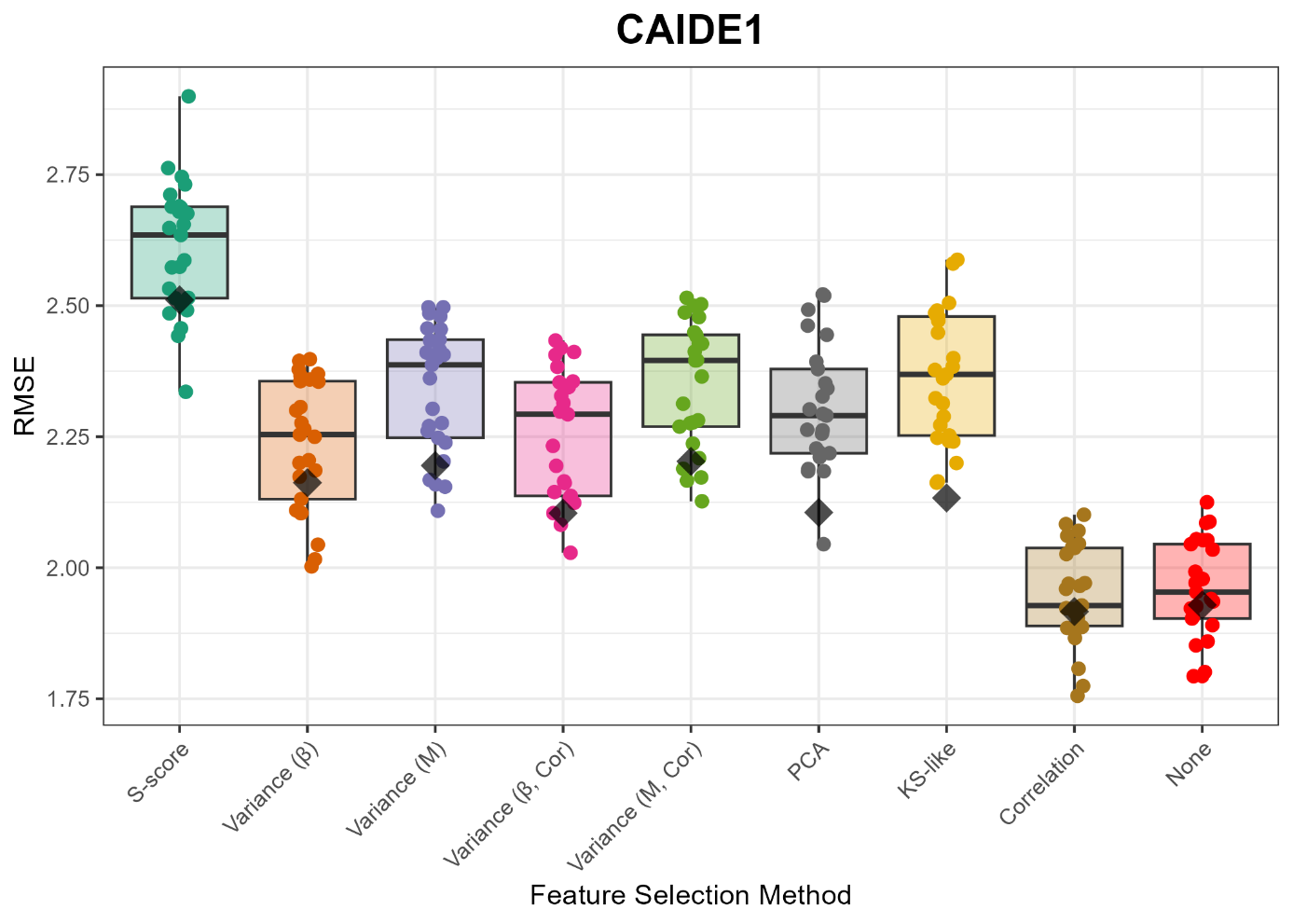

**SUPPLEMENTARY FIGURE 8** Root Mean Squared Error (RMSE) for the prediction of the CAIDE score in the cross-validation (i.e., coloured data points) and test set (i.e., black diamond) for the different feature selection methods.

### Supplementary References

[1] Yengo L, Sidorenko J, Kemper KE, Zheng Z, Wood AR, Weedon MN, et al. Meta-analysis of genome-wide association studies for height and body mass index in∼ 700000 individuals of European ancestry. Hum Mol Genet. 2018;27:3641-9.

[2] Glucose M-Ao, Investigators I-rtC, Consortium GIoAT, Consortium AGENTD, Consortium SATD, Shuldiner AR, et al. Large-scale association analysis provides insights into the genetic architecture and pathophysiology of type 2 diabetes. Nature genetics. 2012;44:981-90.

[3] Marioni RE, Harris SE, Zhang Q, McRae AF, Hagenaars SP, Hill WD, et al. GWAS on family history of Alzheimer’s disease. Translational psychiatry. 2018;8:99.

[4] Willer CJ, Schmidt EM, Sengupta S, Peloso GM, Gustafsson S, Kanoni S, et al. Discovery and refinement of loci associated with lipid levels. Nat Genet. 2013;45:1274-83.

[5] Howard DM, Adams MJ, Clarke T-K, Hafferty JD, Gibson J, Shirali M, et al. Genome-wide meta-analysis of depression identifies 102 independent variants and highlights the importance of the prefrontal brain regions. Nature neuroscience. 2019;22:343-52.

[6] Okbay A, Wu Y, Wang N, Jayashankar H, Bennett M, Nehzati SM, et al. Polygenic prediction of educational attainment within and between families from genome-wide association analyses in 3 million individuals. Nature genetics. 2022;54:437-49.

[7] Nelson CP, Goel A, Butterworth AS, Kanoni S, Webb TR, Marouli E, et al. Association analyses based on false discovery rate implicate new loci for coronary artery disease. Nature genetics. 2017;49:1385-91.

[8] Niarchou M, Byrne EM, Trzaskowski M, Sidorenko J, Kemper KE, McGrath JJ, et al. Genome-wide association study of dietary intake in the UK biobank study and its associations with schizophrenia and other traits. Translational Psychiatry. 2020;10:51.

[9] Abbott L, Bryant S, Churchhouse C, Ganna A, Howrigan D, Palmer D, et al. UK Biobank GWAS round 2. 2018.

[10] Kranzler HR, Zhou H, Kember RL, Vickers Smith R, Justice AC, Damrauer S, et al. Genome-wide association study of alcohol consumption and use disorder in 274,424 individuals from multiple populations. Nature communications. 2019;10:1499.

[11] Wang Z, Emmerich A, Pillon NJ, Moore T, Hemerich D, Cornelis MC, et al. Genome-wide association analyses of physical activity and sedentary behavior provide insights into underlying mechanisms and roles in disease prevention. Nature genetics. 2022;54:1332-44.

[12] Dhana K, Braun KV, Nano J, Voortman T, Demerath EW, Guan W, et al. An epigenome-wide association study of obesity-related traits. American journal of epidemiology. 2018;187:1662-9.

[13] Fraszczyk E, Spijkerman AM, Zhang Y, Brandmaier S, Day FR, Zhou L, et al. Epigenome-wide association study of incident type 2 diabetes: a meta-analysis of five prospective European cohorts. Diabetologia. 2022;65:763-76.

[14] Lohoff FW, Clarke T-K, Kaminsky ZA, Walker RM, Bermingham ML, Jung J, et al. Epigenome-wide association study of alcohol consumption in N= 8161 individuals and relevance to alcohol use disorder pathophysiology: identification of the cystine/glutamate transporter SLC7A11 as a top target. Molecular psychiatry. 2022;27:1754-64.

[15] Braun KV, Dhana K, de Vries PS, Voortman T, van Meurs JB, Uitterlinden AG, et al. Epigenome-wide association study (EWAS) on lipids: the Rotterdam Study. Clinical epigenetics. 2017;9:1-11.

[16] Fernández-Sanlés A, Sayols-Baixeras S, De Moura MC, Esteller M, Subirana I, Torres-Cuevas S, et al. Physical Activity and Genome-wide DNA Methylation: The REGICOR Study. Medicine and science in sports and exercise. 2020;52:589.

[17] Xia Y, Brewer A, Bell JT. DNA methylation signatures of incident coronary heart disease: findings from epigenome-wide association studies. Clinical Epigenetics. 2021;13:1-16.

[18] Li QS, Morrison RL, Turecki G, Drevets WC. Meta-analysis of epigenome-wide association studies of major depressive disorder. Scientific Reports. 2022;12:18361.

[19] Karlsson Linnér R, Marioni RE, Rietveld CA, Simpkin AJ, Davies NM, Watanabe K, et al. An epigenome-wide association study meta-analysis of educational attainment. Molecular psychiatry. 2017;22:1680-90.

[20] Hellbach F, Sinke L, Costeira R, Baumeister S-E, Beekman M, Louca P, et al. Pooled analysis of epigenome-wide association studies of food consumption in KORA, TwinsUK and LLS. European Journal of Nutrition. 2022:1-19.

[21] Richard MA, Huan T, Ligthart S, Gondalia R, Jhun MA, Brody JA, et al. DNA methylation analysis identifies loci for blood pressure regulation. The American Journal of Human Genetics. 2017;101:888-902.

[22] Kennard RW, Stone LA. Computer aided design of experiments. Technometrics. 1969;11:137-48.

[23] Maros ME, Capper D, Jones DT, Hovestadt V, von Deimling A, Pfister SM, et al. Machine learning workflows to estimate class probabilities for precision cancer diagnostics on DNA methylation microarray data. Nature protocols. 2020;15:479-512.

[24] Pidsley R, Y Wong CC, Volta M, Lunnon K, Mill J, Schalkwyk LC. A data-driven approach to preprocessing Illumina 450K methylation array data. BMC genomics. 2013;14:1-10.

[25] Gerritse M. Heterogeneity among trauma-exposed subjects; studied at the multi-omics level. Maastricht: Maastricht University; 2021.
